# Urinary cell mRNA Profiling of Kidney Allograft Recipients: A Systematic Investigation of a Filtration Based Protocol for the Simplification of Urine Processing

**DOI:** 10.1101/2021.06.30.21259736

**Authors:** Catherine Snopkowski, Thalia Salinas, Carol Li, Gabriel Stryjniak, Ruchuang Ding, Vijay Sharma, Manikkam Suthanthiran

## Abstract

**Background:** Kidney transplantation is a life-restorative therapy, but immune rejection undermines allograft survival. Urinary cell mRNA profiles offer a noninvasive means of diagnosing kidney allograft rejection, but urine processing protocols have logistical constraints. We aimed to determine whether the centrifugation-based method for urinary cell mRNA profiling could be replaced with a simpler filtration-based method without undermining quality.

**Methods:** We isolated RNA from urine collected from kidney allograft recipients using the Cornell centrifugation-based protocol (CCBP) or the Zymo filter-based protocol (ZFBP) and compared RNA purity and yield using a spectrophotometer or a fluorometer and measured absolute copy number of transcripts using customized real-time quantitative PCR assays. We investigated the performance characteristics of RNA isolated using ZFBP and stored either at - 80°C or at ambient temperature for 2 to 4 days and also when shipped to our Gene Expression Monitoring (GEM) Core at ambient temperature. We examined the feasibility of initial processing of urine samples by kidney allograft recipients trained by the GEM Core staff and the diagnostic utility for acute rejection, of urine processed using the ZFBP.

**Results:** RNA purity (P=0.0007, Wilcoxon matched paired signed-ranks test) and yield (P<0.0001) were higher with ZFBP vs. CCBP, and absolute copy number of 18S rRNA was similar (P=0.79) following normalization of RNA yield by reverse transcribing a constant amount of RNA isolated using either protocol. RNA purity, yield, and absolute copy numbers of 18S rRNA, TGF-β1 mRNA and microRNA-26a were not different (P>0.05) in the filtrates containing RNA stored either at -80°C or at ambient temperature for 2 to 4 days or shipped overnight at ambient temperature. RNA purity, yield, and absolute copy numbers of 18S rRNA and TGF-β1 mRNA were also not different (P>0.05) between home processed and laboratory processed urine filtrates. Urinary cell levels of mRNA for granzyme B (P=0.01) and perforin (P=0.0002) in the filtrates were diagnostic of acute rejection in human kidney allografts.

**Conclusions:** Urinary cell mRNA profiling was simplified using the ZFBP without undermining RNA quality or diagnostic utility. Home processing by the kidney allograft recipients, the stability of RNA containing filtrates at ambient temperature, and the elimination of the need for centrifuges and freezers represent some of the advantages of ZFBP over the CCBP for urinary cell mRNA profiling.

## INTRODUCTION

Kidney transplantation is a life-saving treatment for those afflicted with end-stage kidney disease, but immune rejection undermines its full benefits (1-3). Accurate diagnosis and prompt treatment of immune rejection represent the best approach for ensuring kidney allograft survival. Percutaneous core needle biopsy is the standard-of-care to diagnose rejection, but this invasive procedure is associated with bleeding, graft failure and even death, and inter-observer variability in the interpretation of biopsies represents another significant challenge (4, 5).

Our laboratory pioneered urinary cell mRNA profiling for the noninvasive diagnosis of immune rejection in human kidney allografts. We customized PCR assays for absolute quantification of mRNA copy number and identified mRNAs diagnostic and prognostic of acute rejection (AR), mRNAs associated with interstitial fibrosis and tubular atrophy (IFTA), and BKV-VP-1 mRNA for the diagnosis of BK virus nephropathy (6-18). Our single center studies paved the way for the multicenter Clinical Trials in Organ Transplantation-04 (CTOT-04) study of 485 prospectively enrolled kidney transplant recipients (ClinicalTrials.gov Identifier: NCT00337220) (15). A total of 4300 urine specimens were collected at 5 academic transplant sites and a Cornell developed urine processing protocol was used to perform initial processing of urine at the collection sites. The protocol included centrifugation of urine to prepare urinary cell pellets, addition of RNAlater to the cell pellets (7), storage at -80°C, and shipment using dry ice (CTOT-04 Laboratory Manual, version 3-0) to our Gene Expression Monitoring (GEM) Core at Weill Cornell. The Core isolated RNA and measured absolute levels of a pre-specified panel of mRNAs using customized RT-qPCR assays developed at the Core.

The CTOT-04 study, in addition to validating previously discovered biomarkers of acute rejection, identified a parsimonious signature diagnostic and prognostic of acute cellular rejection (15). However, the multiple steps involved in urine processing and shipment to the Core imposed logistical challenges for the transplant sites performing the initial processing steps of the collected urine samples. To ensure quality, the transplant coordinators/ laboratory staff from the participating sites received in-person training at the GEM Core prior to the initiation of the CTOT-04 study.

Development of a urine processing protocol that kidney transplant recipients can perform at home has many obvious advantages and should eliminate the need for a centrifuge, a -80°C freezer and visits to the clinic/laboratory for urine collection. A filtration-based method for capturing urinary cells provides an alternative to centrifugation of urine to sediment of urinary cells (19-20). In this investigation, we compared our Cornell centrifugation-based protocol (CCBP) to a commercially available Zymo filter-based protocol (ZFBP) and systematically investigated the various components towards simplification of urine processing.

## MATERIALS & METHODS

### 2.1. Objectives

Our primary objectives were: (i) to compare RNA quality and quantity and transcript abundance in RNA isolated using CCBP or ZFBP; (ii) to determine the RNA quality and quantity in urinary filtrates collected using the ZFBP and stored at ambient temperature and also shipped at ambient temperature; (iii) to determine the feasibility of training kidney allograft recipients to use the filter at home and collect filtrate containing RNA; and (iv) to evaluate whether noninvasive diagnosis of acute rejection is feasible with the filtrate collected using ZFBP.

### 2.2. Research Protocols

Urine samples were collected from kidney allograft recipients managed at our transplant center. The samples were processed using either CCBP (15) or ZFBP. In the ZFBP, RNA was isolated from urine using Urine RNA Isolation Kit™ purchased from Zymo Research, Orange, CA. The kit (Product R 1038 and R1039) included RNA extraction buffer plus, RNA wash buffer, RNA elution buffer, cell collector filter, Zymo-Spin I column, and collection tubes. The initial version was later modified by the manufacturer, and the later version termed ZR Urine Isolation Kit™ was used for the experiments performed under Protocols 2-5, and the modified kit included proprietary urine RNA buffer, RNA prep buffer, RNA wash buffer, DNase/RNase free-water prep buffer, ZRC GF™ filter, Zymo-Spin IC™ columns and collection tubes. We examined whether the ZFBP for RNA isolation is noninferior to our published CCBP for RNA isolation with respect to RNA purity, RNA yield, and transcript abundance in urine collected from kidney allograft recipients (Protocol 1); whether the performance characteristics of the RNA in the urine filtrate, stored at room temperature for 2 to 4 days, is non-inferior to the filtrate stored at -80°C (Protocol 2); whether the performance characteristics of the RNA in the urine filtrate mailed overnight at ambient temperature to our GEM Core is non-inferior to the filtrate stored at -80°C (Protocol 3); whether the performance characteristics of the RNA in the urine filtrate collected by the kidney allograft recipients at home is non-inferior to the RNA in the urine filtrate prepared by the GEM Core Staff (Protocol 4); and whether noninvasive diagnosis of acute rejection in the human kidney allograft is feasible using the ZFBP (Protocol 5).

The study participants provided written informed consent for participation in the noninvasive diagnosis of kidney allograft rejection study and Weill Cornell Medicine Institutional Review Board approved urine collection from kidney allograft recipients for gene expression profiling.

### 2.3. RNA Characteristics

We used NanoDrop® ND-1000 spectrophotometer (ThermoFisher Scientific, Waltham, MA) to measure the concentration of the RNA isolated using the CCBP or ZFBP (absorbance at 260nm) and RNA purity (the ratio of absorbance at 260 and 280). Qubit Fluorometer (ThermoFisher Scientific) was used to measure the concentration of RNA extracted from the urine specimens in select instances. We used identical protocols for the reverse transcription of RNA isolated using either CCBP or ZFBP, as previously described (15). In brief, total RNA was reverse transcribed to cDNA using the TaqMan reverse transcription kit (Cat. N808-0234, Applied Biosystems). We normalized RNA yield from different urine specimens by performing reverse transcription after adjustment of isolated RNA to a concentration of 1.0μg of RNA in 100 μl volume. The reverse transcription reaction contained the final concentration of 1x TaqMan RT buffer, 5.5 mM of Magnesium Chloride, 500 μM each of 4 dNTPs, 2.5 μM of Random Hexamer, 0.4 Unit/μl of RNase inhibitor, and 1.25 Unit/μl of MultiScribe Reverse Transcriptase. The sample was incubated at 25°C for 10 min, 48°C for 30 min, and 95°C for 5 min.

The absolute copy number of the reference gene 18S rRNA and the absolute copy number of a constitutively expressed TGF-β1 mRNA were measured using real-time quantitative PCR assays customized in our Core by the incorporation of in-house generated Bak amplicon (customized RT-qPCR assay) to calculate absolute copy numbers of the transcripts (15). The diagnosis of acute rejection and the absence of acute or chronic rejection were confirmed in each instance by percutaneous core needle biopsy of the kidney allograft, and the biopsies were classified by the pathologist masked to urine mRNA profiling data and using the Banff biopsy classification schema (21). Urinary cell levels of mRNA for perforin and granzyme B, validated biomarkers of acute rejection (15), were measured using gene specific oligonucleotide primer pairs and gene specific TaqMan probes in the customized RT-qPCR assays. The primers and probes used to measure absolute copy number of 18S rRNA, TGF-β1 mRNA, perforin mRNA and granzyme B mRNA were designed in our GEM Core and the sequences of the primers and probes have been published (15).

### 2.4. Statistical Analysis

Absolute copy numbers of transcripts were calculated based on Bak amplicon based standard curve in the customized RT-qPCR assays, as reported (15). Median, 25^th^ percentile and 75^th^ percentile values were calculated for continuous variables. Wilcoxon matched paired signed-rank test or Mann-Whitney test was used for statistical analysis for continuous variables. A P value <0.05 was considered significant. We used GraphPad Prism 9 statistical software package for data analysis.

## 3. RESULTS

### 3.1. RNA Purity, Yield, and Transcript Abundance: CCBP vs. ZFBP

In Protocol 1, we compared RNA purity and yield and the absolute copy number of a reference gene 18S rRNA in 47 urine samples collected from 39 unique kidney allograft recipients and processed using either the CCBP (15) or the ZFBP. Figure 1 shows the protocol for processing the urine specimens. Each urine sample from an individual patient was divided into 2 aliquots prior to processing. In the CCBP, urine samples were centrifuged at 2000g for 30 minutes at ambient temperature for the sedimentation of urinary cells and the urine cell pellets were then processed as outlined in Figure 1. The centrifugation step was performed within 4 to 6 hours of urine collection because urine is rich in RNA hydrolyzing enzymes (22). In the ZFBP, urine was pushed through the Zymo filter using a syringe and the urinary cells trapped by the filter were lysed using the RNA extraction buffer plus included in the kit and RNA containing filtrate was collected. The processing of RNA containing filtrate was also performed within 4 to 6 hours of urine collection. The cell pellet prepared using the CCBP and the RNA containing filtrate prepared using the ZFBP were then stored at -80°C prior to RNA isolation. Following retrieval of the cell pellets and the filtrates from storage, Rneasy Mini Kit™ was used for the isolation of RNA from urinary cell pellets, and the Zymo-Spin ™ IC column was used for RNA isolation from the filtrate. The purity and the amount of RNA isolated by either protocol were measured and RNA yield normalized by reverse transcription of RNA to cDNA at a concentration of 1.0 microgram in 100 μl.

**Figure 1.**
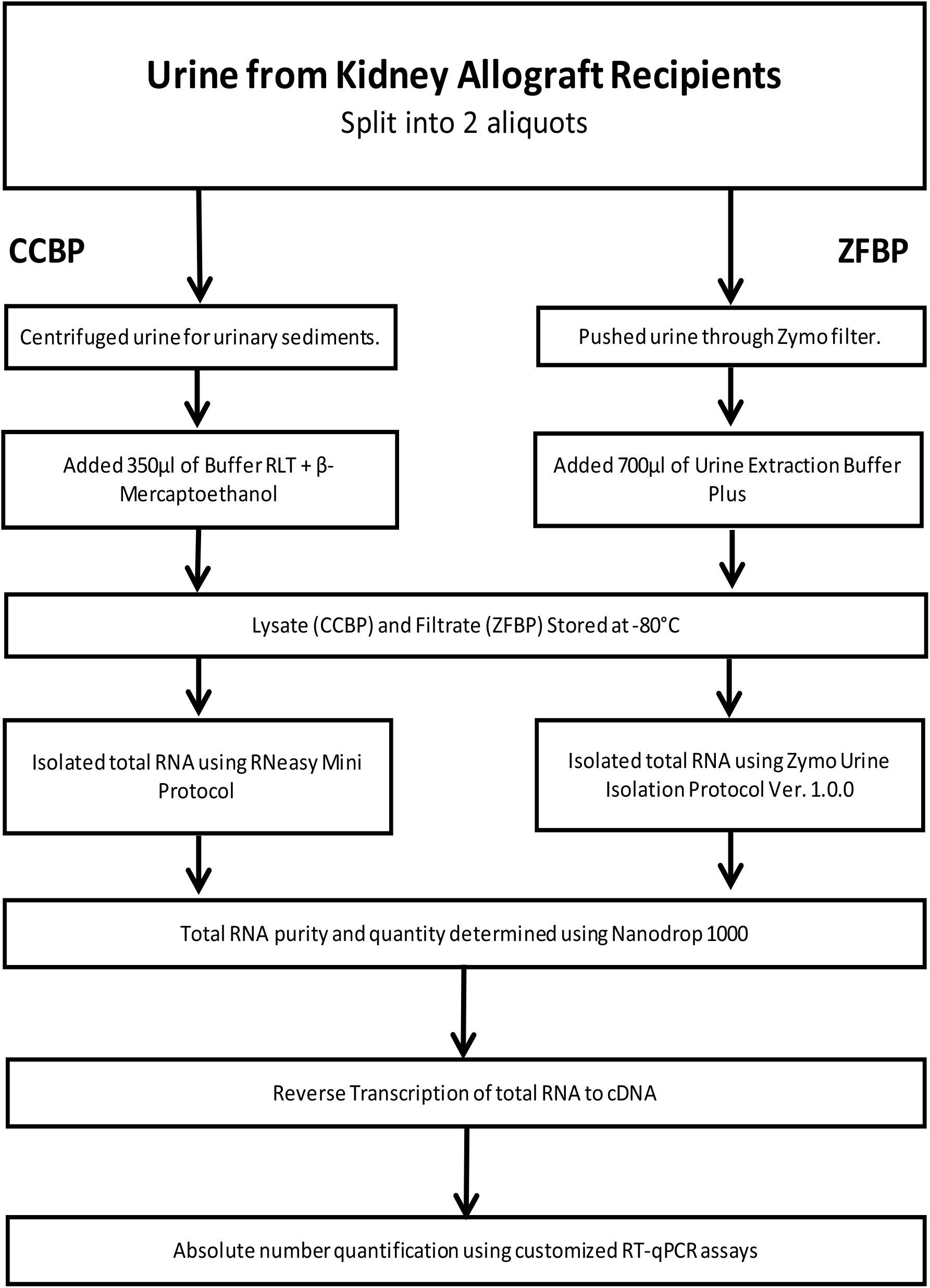
Flow chart for comparing Cornell Centrifugation Based Protocol (CCBP) vs. Zymo Filter Based Protocol (ZFBP). Forty-seven consecutive urine samples collected from 39 unique kidney allograft recipients were divided into 2 aliquots and processed using either the CCBP or the ZFBP. In the CCBP, urine was centrifuged at 2000g for 30 minutes at room temperature for the sedimentation of urinary cells and lysed with Buffer RLT + β- Mercaptoethanol. In the ZFBP, urine was pushed through the Zymo cell collection filter and 700μl of RNA Extraction Buffer Plus was pushed through the filter to collect urinary cell filtrate containing RNA. RNA was isolated from the lysates using RNeasy Mini Kit (Qiagen, Cat. 74104) and RNA was isolated from urine cell filtrate using Urine RNA Isolation Kit (Zymo, Cat. R1038/1039 Ver. 1.0.0) where the elution step was modified to include 25μl TE and 25μl RNAse-free water, and an additional minute of incubation in between elution with TE and RNAse-free water. Total RNA was measured using the Nanodrop 1000 Spectrophotometer. Nanodrop measurement replicates were within 1ng/μl concentration and 0.2 for the A260/ 280 ratio. Total RNA was reverse transcribed based on the Nanodrop 1000 readings to cDNA using TaqMan reverse transcription kit (Applied Biosystems, Cat. N8080234). Absolute copy number of 18S rRNA was measured using 18S rRNA specific oligonucleotide primer pair and 18S rRNA specific TaqMan probe in the customized RT-qPCR assay on an ABI 7700 Detection System (Applied Biosystems).

Data from RNA isolated from the 47 urine samples processed using the CCBP or the ZFBP are shown in Table 1A and the demographics of the study cohort are summarized in Table 1B. Among these 47 samples, equal volumes of urine were processed in 38 instances and unequal volumes in the remaining 9 instances. The median (25^th^ percentile and 75^th^ percentile) ratio of A260 to A280 for the RNA isolated from all 47 paired samples using the CCBP was 1.96 (1.56 and 2.08) and 2.00 (1.94 and 2.07) with the ZFBP (Mann Whitney P=0.06). RNA yield from all 47 paired urine samples was 0.30 (0.10 and 0.70) micrograms with the CCBP and 0.81 (0.33 and 2.40) micrograms with the ZFBP (P<0.0001). The higher yield of total RNA with the ZFBP as compared to the CCBP was also observed in an analysis restricted to the 38 paired urine samples of equal volumes; 0.15 (0.10 and 0.44) micrograms with the CCBP and 0.47 (0.30 and 1.33) micrograms with the ZFBP (Wilcoxon matched pair signed-ranks test P<0.0001).

**Table 1A.**
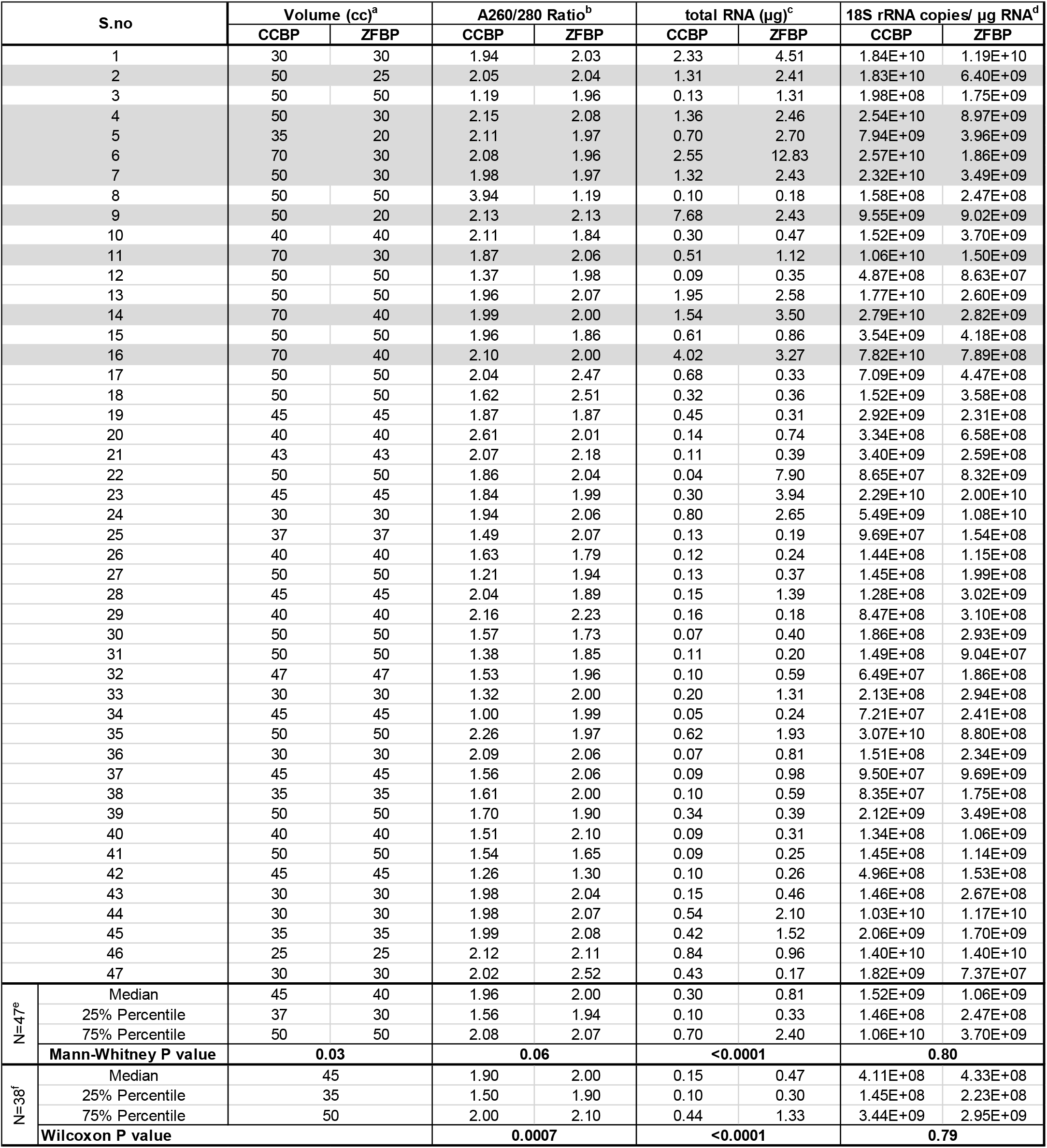
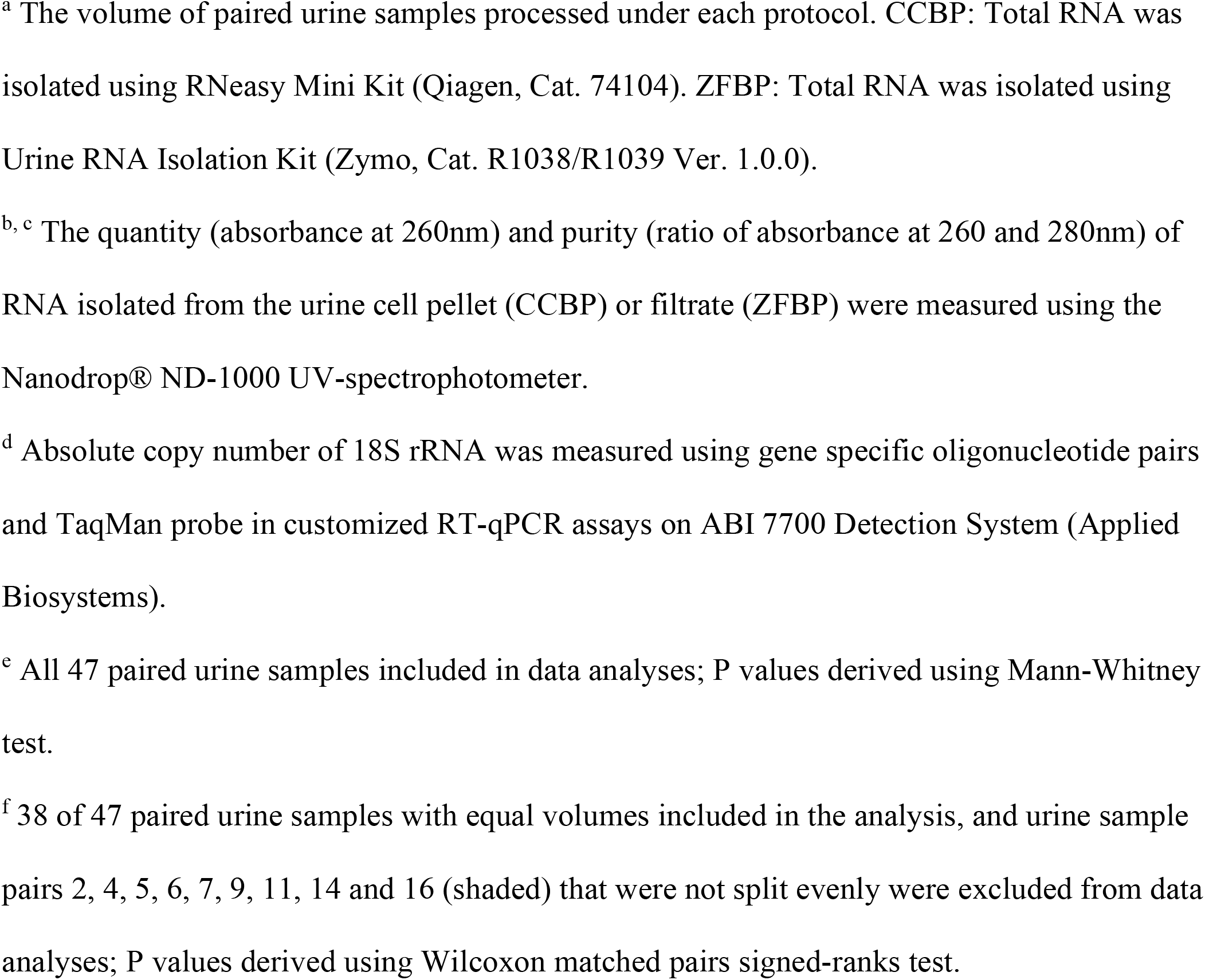
RNA purity, yield, and absolute copy number of 18S rRNA in urine samples processed using the CCBP or the ZFBP.

| S.no |  | Volume (cc)<sup>a</sup> |  | A260/280 Ratio<sup>b</sup> |  | total RNA (µg)<sup>c</sup> |  | 18S rRNA copies/ µg RNA<sup>d</sup> |  |
| --- | --- | --- | --- | --- | --- | --- | --- | --- | --- |
|  |  | CCBP | ZFBP | CCBP | ZFBP | CCBP | ZFBP | CCBP | ZFBP |
| 1 |  | 30 | 30 | 1.94 | 2.03 | 2.33 | 4.51 | 1.84E+10 | 1.19E+10 |
| 2 |  | 50 | 25 | 2.05 | 2.04 | 1.31 | 2.41 | 1.83E+10 | 6.40E+09 |
| 3 |  | 50 | 50 | 1.19 | 1.96 | 0.13 | 1.31 | 1.98E+08 | 1.75E+09 |
| 4 |  | 50 | 30 | 2.15 | 2.08 | 1.36 | 2.46 | 2.54E+10 | 8.97E+09 |
| 5 |  | 35 | 20 | 2.11 | 1.97 | 0.70 | 2.70 | 7.94E+09 | 3.96E+09 |
| 6 |  | 70 | 30 | 2.08 | 1.96 | 2.55 | 12.83 | 2.57E+10 | 1.86E+09 |
| 7 |  | 50 | 30 | 1.98 | 1.97 | 1.32 | 2.43 | 2.32E+10 | 3.49E+09 |
| 8 |  | 50 | 50 | 3.94 | 1.19 | 0.10 | 0.18 | 1.58E+08 | 2.47E+08 |
| 9 |  | 50 | 20 | 2.13 | 2.13 | 7.68 | 2.43 | 9.55E+09 | 9.02E+09 |
| 10 |  | 40 | 40 | 2.11 | 1.84 | 0.30 | 0.47 | 1.52E+09 | 3.70E+09 |
| 11 |  | 70 | 30 | 1.87 | 2.06 | 0.51 | 1.12 | 1.06E+10 | 1.50E+09 |
| 12 |  | 50 | 50 | 1.37 | 1.98 | 0.09 | 0.35 | 4.87E+08 | 8.63E+07 |
| 13 |  | 50 | 50 | 1.96 | 2.07 | 1.95 | 2.58 | 1.77E+10 | 2.60E+09 |
| 14 |  | 70 | 40 | 1.99 | 2.00 | 1.54 | 3.50 | 2.79E+10 | 2.82E+09 |
| 15 |  | 50 | 50 | 1.96 | 1.86 | 0.61 | 0.86 | 3.54E+09 | 4.18E+08 |
| 16 |  | 70 | 40 | 2.10 | 2.00 | 4.02 | 3.27 | 7.82E+10 | 7.89E+08 |
| 17 |  | 50 | 50 | 2.04 | 2.47 | 0.68 | 0.33 | 7.09E+09 | 4.47E+08 |
| 18 |  | 50 | 50 | 1.62 | 2.51 | 0.32 | 0.36 | 1.52E+09 | 3.58E+08 |
| 19 |  | 45 | 45 | 1.87 | 1.87 | 0.45 | 0.31 | 2.92E+09 | 2.31E+08 |
| 20 |  | 40 | 40 | 2.61 | 2.01 | 0.14 | 0.74 | 3.34E+08 | 6.58E+08 |
| 21 |  | 43 | 43 | 2.07 | 2.18 | 0.11 | 0.39 | 3.40E+09 | 2.59E+08 |
| 22 |  | 50 | 50 | 1.86 | 2.04 | 0.04 | 7.90 | 8.65E+07 | 8.32E+09 |
| 23 |  | 45 | 45 | 1.84 | 1.99 | 0.30 | 3.94 | 2.29E+10 | 2.00E+10 |
| 24 |  | 30 | 30 | 1.94 | 2.06 | 0.80 | 2.65 | 5.49E+09 | 1.08E+10 |
| 25 |  | 37 | 37 | 1.49 | 2.07 | 0.13 | 0.19 | 9.69E+07 | 1.54E+08 |
| 26 |  | 40 | 40 | 1.63 | 1.79 | 0.12 | 0.24 | 1.44E+08 | 1.15E+08 |
| 27 |  | 50 | 50 | 1.21 | 1.94 | 0.13 | 0.37 | 1.45E+08 | 1.99E+08 |
| 28 |  | 45 | 45 | 2.04 | 1.89 | 0.15 | 1.39 | 1.28E+08 | 3.02E+09 |
| 29 |  | 40 | 40 | 2.16 | 2.23 | 0.16 | 0.18 | 8.47E+08 | 3.10E+08 |
| 30 |  | 50 | 50 | 1.57 | 1.73 | 0.07 | 0.40 | 1.86E+08 | 2.93E+09 |
| 31 |  | 50 | 50 | 1.38 | 1.85 | 0.11 | 0.20 | 1.49E+08 | 9.04E+07 |
| 32 |  | 47 | 47 | 1.53 | 1.96 | 0.10 | 0.59 | 6.49E+07 | 1.86E+08 |
| 33 |  | 30 | 30 | 1.32 | 2.00 | 0.20 | 1.31 | 2.13E+08 | 2.94E+08 |
| 34 |  | 45 | 45 | 1.00 | 1.99 | 0.05 | 0.24 | 7.21E+07 | 2.41E+08 |
| 35 |  | 50 | 50 | 2.26 | 1.97 | 0.62 | 1.93 | 3.07E+10 | 8.80E+08 |
| 36 |  | 30 | 30 | 2.09 | 2.06 | 0.07 | 0.81 | 1.51E+08 | 2.34E+09 |
| 37 |  | 45 | 45 | 1.56 | 2.06 | 0.09 | 0.98 | 9.50E+07 | 9.69E+09 |
| 38 |  | 35 | 35 | 1.61 | 2.00 | 0.10 | 0.59 | 8.35E+07 | 1.75E+08 |
| 39 |  | 50 | 50 | 1.70 | 1.90 | 0.34 | 0.39 | 2.12E+09 | 3.49E+08 |
| 40 |  | 40 | 40 | 1.51 | 2.10 | 0.09 | 0.31 | 1.34E+08 | 1.06E+09 |
| 41 |  | 50 | 50 | 1.54 | 1.65 | 0.09 | 0.25 | 1.45E+08 | 1.14E+09 |
| 42 |  | 45 | 45 | 1.26 | 1.30 | 0.10 | 0.26 | 4.96E+08 | 1.53E+08 |
| 43 |  | 30 | 30 | 1.98 | 2.04 | 0.15 | 0.46 | 1.46E+08 | 2.67E+08 |
| 44 |  | 30 | 30 | 1.98 | 2.07 | 0.54 | 2.10 | 1.03E+10 | 1.17E+10 |
| 45 |  | 35 | 35 | 1.99 | 2.08 | 0.42 | 1.52 | 2.06E+09 | 1.70E+09 |
| 46 |  | 25 | 25 | 2.12 | 2.11 | 0.84 | 0.96 | 1.40E+10 | 1.40E+10 |
| 47 |  | 30 | 30 | 2.02 | 2.52 | 0.43 | 0.17 | 1.82E+09 | 7.37E+07 |
| N=47<sup>e</sup> | Median | 45 | 40 | 1.96 | 2.00 | 0.30 | 0.81 | 1.52E+09 | 1.06E+09 |
|  | 25% Percentile | 37 | 30 | 1.56 | 1.94 | 0.10 | 0.33 | 1.46E+08 | 2.47E+08 |
|  | 75% Percentile | 50 | 50 | 2.08 | 2.07 | 0.70 | 2.40 | 1.06E+10 | 3.70E+09 |
|  | Mann-Whitney P value | 0.03 |  | 0.06 |  | <0.0001 |  | 0.80 |  |
| N=38<sup>f</sup> | Median | 45 |  | 1.90 | 2.00 | 0.15 | 0.47 | 4.11E+08 | 4.33E+08 |
|  | 25% Percentile | 35 |  | 1.50 | 1.90 | 0.10 | 0.30 | 1.45E+08 | 2.23E+08 |
|  | 75% Percentile | 50 |  | 2.00 | 2.10 | 0.44 | 1.33 | 3.44E+09 | 2.95E+09 |
|  | Wilcoxon P value |  |  | 0.0007 |  | <0.0001 |  | 0.79 |  |

<sup>a</sup> The volume of paired urine samples processed under each protocol. CCBP: Total RNA was isolated using RNeasy Mini Kit (Qiagen, Cat. 74104). ZFBP: Total RNA was isolated using Urine RNA Isolation Kit (Zymo, Cat. R1038/R1039 Ver. 1.0.0).
<sup>b, c</sup> The quantity (absorbance at 260nm) and purity (ratio of absorbance at 260 and 280nm) of RNA isolated from the urine cell pellet (CCBP) or filtrate (ZFBP) were measured using the Nanodrop® ND-1000 UV-spectrophotometer.
<sup>d</sup> Absolute copy number of 18S rRNA was measured using gene specific oligonucleotide pairs and TaqMan probe in customized RT-qPCR assays on ABI 7700 Detection System (Applied Biosystems).
<sup>e</sup> All 47 paired urine samples included in data analyses; P values derived using Mann-Whitney test.
<sup>f</sup> 38 of 47 paired urine samples with equal volumes included in the analysis, and urine sample pairs 2, 4, 5, 6, 7, 9, 11, 14 and 16 (shaded) that were not split evenly were excluded from data analyses; P values derived using Wilcoxon matched pairs signed-ranks test.

**Table 1B.** Characteristics of study cohort.

| **Characteristics of Kidney Allograft Recipients** |  |
| --- | --- |
| Number of patients | 39 |
| Age, Years, Median (Min- Max) | 46 (22-80) |
| Sex, Male / Female | 20 / 19 |
| Time from Transplantation to Urine Collection, Median (Min-Max) | 12 months<br>(23 days-78 months) |
| Type of Donor Kidney, Deceased / Living Related / Living Unrelated | 25 / 11 / 3 |

We normalized RNA yield from different urine samples by reverse transcribing the RNA isolated by either the CCBP or the ZFBP at a concentration of 1.0 microgram in 100 microliters, as previously described (15). Absolute number of 18S rRNA copy number was quantified using 18S rRNA specific oligonucleotide primer pairs and 18S rRNA specific TaqMan probe in the customized RT-qPCR assays. We previously established 5×10^7^ 18S rRNA copy numbers per microgram of total RNA as one of the quality thresholds for RNA adequacy (15). RNA from all 47 urine specimens, isolated with either protocol, met the 5×10^7^ 18S rRNA copy numbers per microgram of RNA threshold, and the copy number found in each urine sample is shown in Table 1. The absolute copy number of the reference gene 18S rRNA in the 47 urine specimens from 39 unique kidney allograft recipients was 1.52 E+09 (1.46E+08 and 1.06E+10) copies per microgram of total RNA with the CCBP and 1.06 E+09 (2.47E+08 and 3.70E+09) copies per microgram of total RNA in the paired urine samples with the ZFBP (P= 0.80).

Data provided in Table 1 demonstrate that RNA purity and RNA yield are higher with the ZFBP compared to CCBP and that 18S rRNA copy number is not different following normalization of RNA yield at the reverse transcription step.

### 3.2. Urinary Cell Profiling with Filtrates Stored at Ambient Temperature

In the multicenter CTOT-04 study, urine was centrifuged to prepare urine cell pellets at each participating site, RNAlater was added to the cell pellet (7) and the pellets were stored at -80°C at the collection sites and then shipped in batches in cold (dry ice) containers to the GEM Core (15). Because of the high content of RNA hydrolyzing enzymes in urine (22), the urine samples collected from the kidney allograft recipients were processed within 4 to 6 hours of collection. The need for -80°C freezer to store the cell pellets and the need for special packaging for shipment (e.g., dry ice) represent logistical challenges. Having identified that the ZFBP is noninferior to CCBP and may even be superior in terms of RNA purity and yield (Table 1), we investigated whether absolute copy number of transcripts is similar between the ZFBP filtrates stored for 2 or 4 days at either -80°C or at ambient temperature using the protocol illustrated in Figure 2.

**Figure 2.**
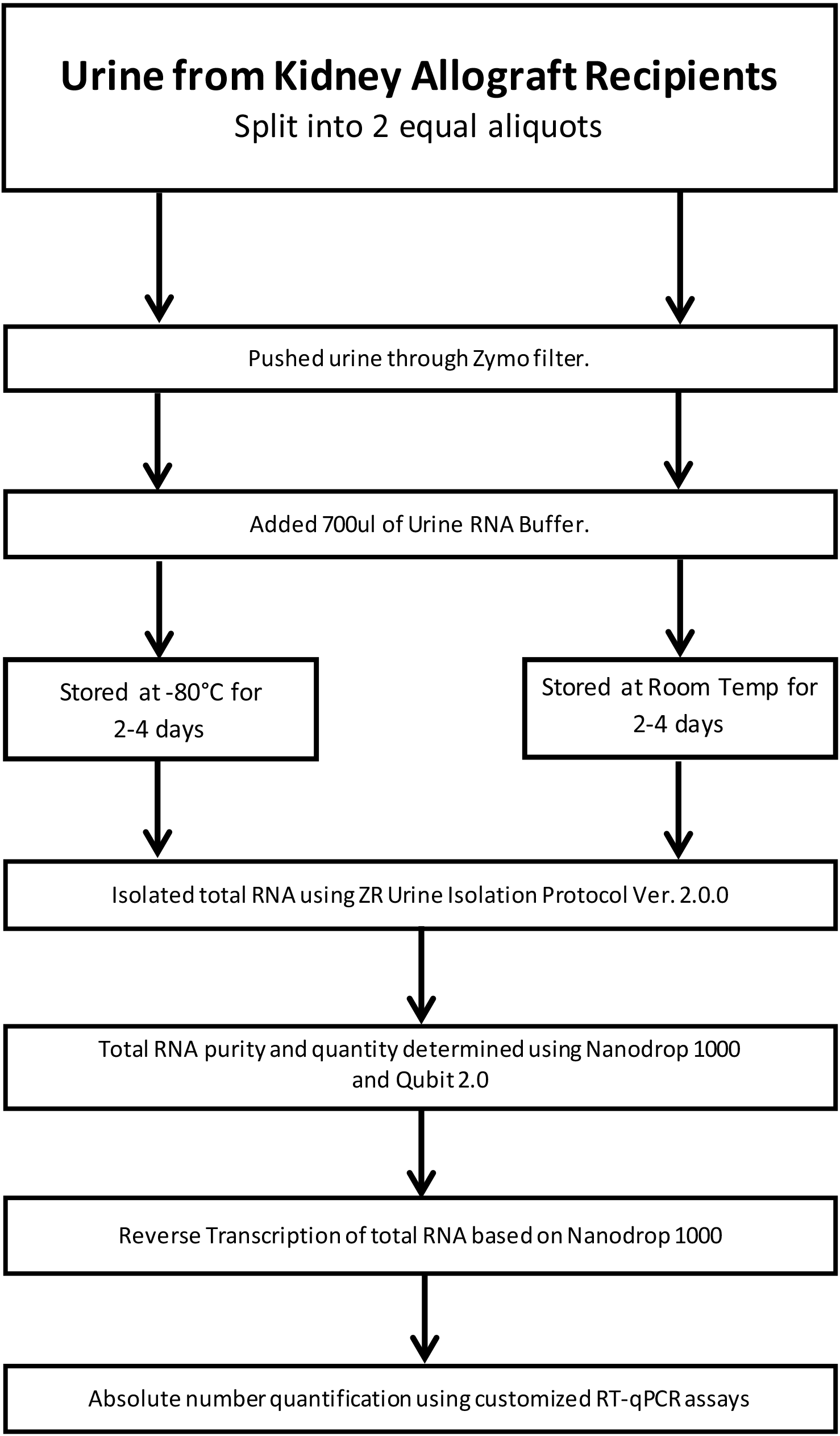
Flow chart for the evaluation of RNA containing filtrates stored either at -80°C or Ambient (Room) Temperature. Urine samples collected from kidney allograft recipients were divided into 2 equal aliquots and processed using the ZR Urine RNA Isolation Kit™. Urine was pushed through the ZRC GF™ filter and 700μl of urine RNA buffer was added to the filter to collect urinary cell filtrate containing total RNA. The filtrates were stored either at -80°C or at room temperature for 2 or 4 days. The amount of RNA isolated from the filtrates was measured using the Nanodrop 1000 Spectrophotometer and Qubit 2.0 fluorometer. The Nanodrop measurement replicates were within 1ng/μl concentration and 0.2 for the A260/280 ratio. For the measurement of 18S rRNA and TGF-β1 mRNA, RNA was reverse transcribed to cDNA using TaqMan reverse transcription kit. For the measurement of miR-26a, total RNA was reverse transcribed using miR-26a -specific TaqMan MicroRNA reverse transcription kit. Absolute copy numbers of 18S rRNA, TGF-β1 mRNA, and hsa-miR-26a miRNA were measured using customized Rt-qPCR assays using gene specific oligonucleotide pairs and TaqMan probes on 7500 Fast Real-Time PCR System.

Among the 12 urine samples from 12 kidney allograft recipients, one urine sample did not yield sufficient RNA for reverse transcription. Table 2A shows data from each filtrate and Table 2B is a summary of the study cohort. The median (25^th^ percentile and 75^th^ percentile) A260/280 ratio of RNA isolated from the 11 filtrates stored at -80°C for 2 or 4 days prior to RNA isolation was 1.87 (1.70 and 1.96) and 1.80 (1.59 and 1.94) in the paired 11 filtrates stored at ambient temperature for 2 or 4 days prior to RNA isolation (Wilcoxon matched-pairs signed-rank test P=0.57). The amount of RNA extracted from the 11 filtrates stored at -80°C was 0.43 (0.21 and 1.30) micrograms and 0.48 (0.24 and 1.38) micrograms of RNA from the paired 11 filtrates stored at ambient temperature (P=0.04) (Table 2A). These measurements were made using the NanoDrop spectrophotometer. We also measured the amount of RNA extracted from the filtrates using the Qubit fluorometer considered to be more sensitive and specific for RNA estimation compared to spectrophotometry-based quantification. The amount of RNA, measured using the Qubit Fluorometer, was lower compared to the RNA quantity measured with the NanoDrop spectrophotometer, and was 0.23 (0.14 and 0.74) micrograms of total RNA from the 11 filtrates stored at -80°C and 0.20 (0.15 and 0.74) micrograms in the paired 11 filtrates stored at ambient temperature (P=0.30) (Table 2A).

**Table 2A.**
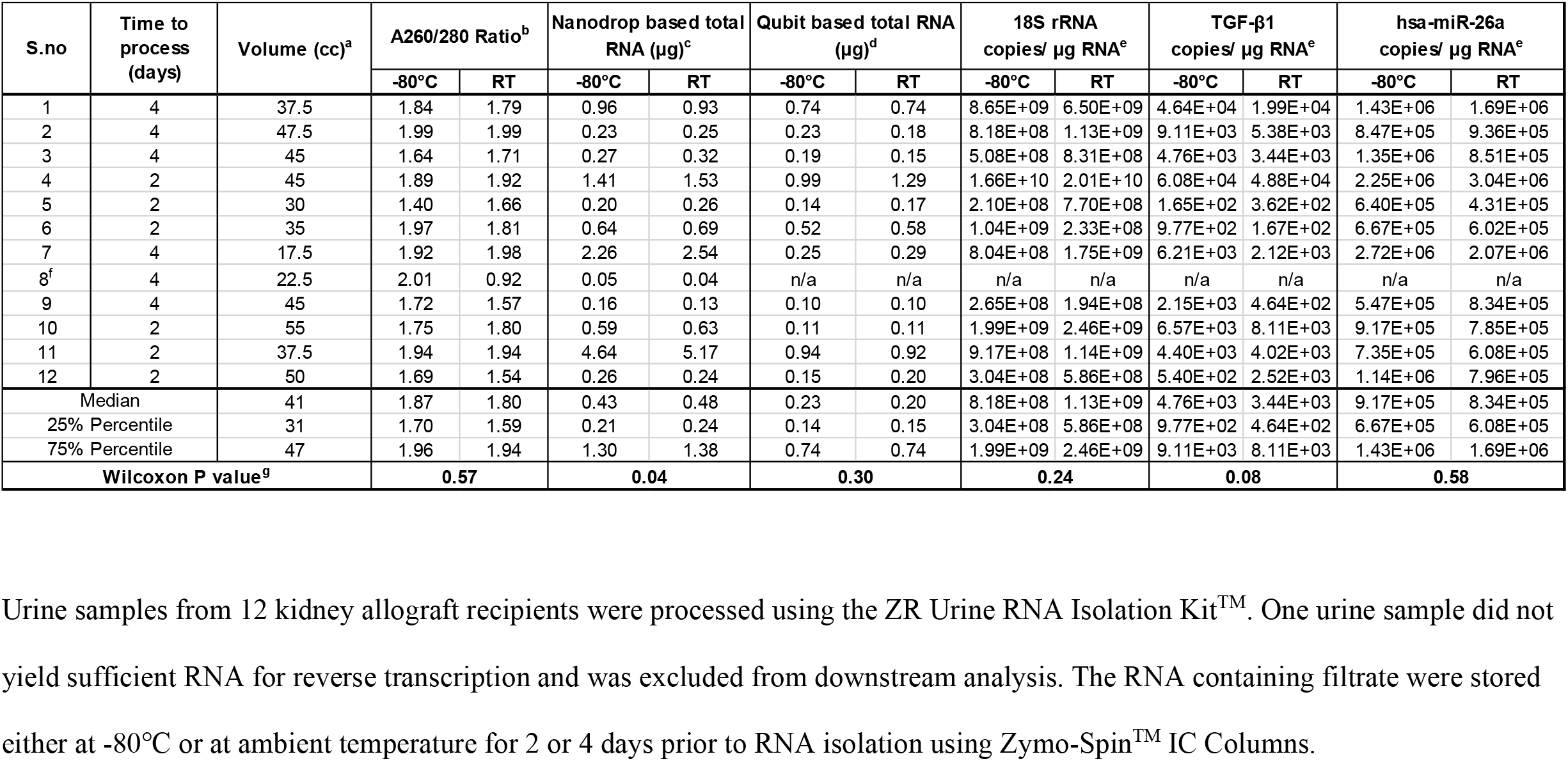
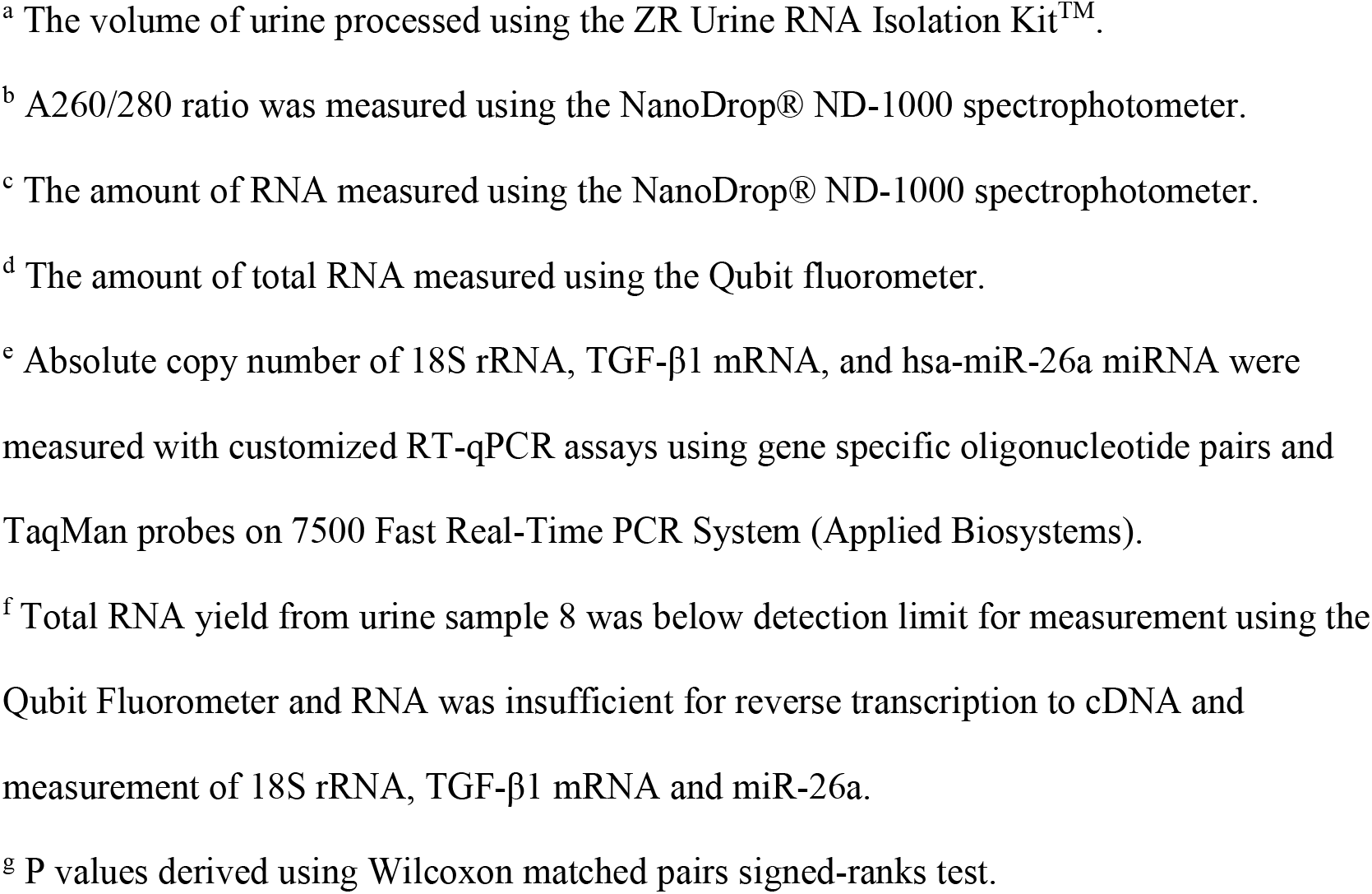
Performance characteristics of total RNA isolated from the filtrates stored either at -80°C or Ambient (Room) Temperature.

| S.no | Time to process (days) | Volume (cc)<sup>a</sup> | A260/280 Ratio<sup>b</sup> |  | Nanodrop based total RNA (µg)<sup>c</sup> |  | Qubit based total RNA (µg)<sup>d</sup> |  | 18S rRNA copies/ µg RNA<sup>e</sup> |  | TGF-β1 copies/ µg RNA<sup>e</sup> |  | hsa-miR-26a copies/ µg RNA<sup>e</sup> |  |
| --- | --- | --- | --- | --- | --- | --- | --- | --- | --- | --- | --- | --- | --- | --- |
|  |  |  | -80°C | RT | -80°C | RT | -80°C | RT | -80°C | RT | -80°C | RT | -80°C | RT |
| 1 | 4 | 37.5 | 1.84 | 1.79 | 0.96 | 0.93 | 0.74 | 0.74 | 8.65E+09 | 6.50E+09 | 4.64E+04 | 1.99E+04 | 1.43E+06 | 1.69E+06 |
| 2 | 4 | 47.5 | 1.99 | 1.99 | 0.23 | 0.25 | 0.23 | 0.18 | 8.18E+08 | 1.13E+09 | 9.11E+03 | 5.38E+03 | 8.47E+05 | 9.36E+05 |
| 3 | 4 | 45 | 1.64 | 1.71 | 0.27 | 0.32 | 0.19 | 0.15 | 5.08E+08 | 8.31E+08 | 4.76E+03 | 3.44E+03 | 1.35E+06 | 8.51E+05 |
| 4 | 2 | 45 | 1.89 | 1.92 | 1.41 | 1.53 | 0.99 | 1.29 | 1.66E+10 | 2.01E+10 | 6.08E+04 | 4.88E+04 | 2.25E+06 | 3.04E+06 |
| 5 | 2 | 30 | 1.40 | 1.66 | 0.20 | 0.26 | 0.14 | 0.17 | 2.10E+08 | 7.70E+08 | 1.65E+02 | 3.62E+02 | 6.40E+05 | 4.31E+05 |
| 6 | 2 | 35 | 1.97 | 1.81 | 0.64 | 0.69 | 0.52 | 0.58 | 1.04E+09 | 2.33E+08 | 9.77E+02 | 1.67E+02 | 6.67E+05 | 6.02E+05 |
| 7 | 4 | 17.5 | 1.92 | 1.98 | 2.26 | 2.54 | 0.25 | 0.29 | 8.04E+08 | 1.75E+09 | 6.21E+03 | 2.12E+03 | 2.72E+06 | 2.07E+06 |
| 8<sup>f</sup> | 4 | 22.5 | 2.01 | 0.92 | 0.05 | 0.04 | n/a | n/a | n/a | n/a | n/a | n/a | n/a | n/a |
| 9 | 4 | 45 | 1.72 | 1.57 | 0.16 | 0.13 | 0.10 | 0.10 | 2.65E+08 | 1.94E+08 | 2.15E+03 | 4.64E+02 | 5.47E+05 | 8.34E+05 |
| 10 | 2 | 55 | 1.75 | 1.80 | 0.59 | 0.63 | 0.11 | 0.11 | 1.99E+09 | 2.46E+09 | 6.57E+03 | 8.11E+03 | 9.17E+05 | 7.85E+05 |
| 11 | 2 | 37.5 | 1.94 | 1.94 | 4.64 | 5.17 | 0.94 | 0.92 | 9.17E+08 | 1.14E+09 | 4.40E+03 | 4.02E+03 | 7.35E+05 | 6.08E+05 |
| 12 | 2 | 50 | 1.69 | 1.54 | 0.26 | 0.24 | 0.15 | 0.20 | 3.04E+08 | 5.86E+08 | 5.40E+02 | 2.52E+03 | 1.14E+06 | 7.96E+05 |
| Median |  | 41 | 1.87 | 1.80 | 0.43 | 0.48 | 0.23 | 0.20 | 8.18E+08 | 1.13E+09 | 4.76E+03 | 3.44E+03 | 9.17E+05 | 8.34E+05 |
| 25% Percentile |  | 31 | 1.70 | 1.59 | 0.21 | 0.24 | 0.14 | 0.15 | 3.04E+08 | 5.86E+08 | 9.77E+02 | 4.64E+02 | 6.67E+05 | 6.08E+05 |
| 75% Percentile |  | 47 | 1.96 | 1.94 | 1.30 | 1.38 | 0.74 | 0.74 | 1.99E+09 | 2.46E+09 | 9.11E+03 | 8.11E+03 | 1.43E+06 | 1.69E+06 |
| Wilcoxon P value<sup>g</sup> |  |  | 0.57 |  | 0.04 |  | 0.30 |  | 0.24 |  | 0.08 |  | 0.58 |  |

Urine samples from 12 kidney allograft recipients were processed using the ZR Urine RNA Isolation Kit<sup>TM</sup>. One urine sample did not yield sufficient RNA for reverse transcription and was excluded from downstream analysis. The RNA containing filtrate were stored either at -80°C or at ambient temperature for 2 or 4 days prior to RNA isolation using Zymo-Spin<sup>TM</sup> IC Columns.
<sup>a</sup> The volume of urine processed using the ZR Urine RNA Isolation Kit<sup>TM</sup>.
<sup>b</sup> A260/280 ratio was measured using the NanoDrop® ND-1000 spectrophotometer.
<sup>c</sup> The amount of RNA measured using the NanoDrop® ND-1000 spectrophotometer.
<sup>d</sup> The amount of total RNA measured using the Qubit fluorometer.
<sup>e</sup> Absolute copy number of 18S rRNA, TGF-β1 mRNA, and hsa-miR-26a miRNA were measured with customized RT-qPCR assays using gene specific oligonucleotide pairs and TaqMan probes on 7500 Fast Real-Time PCR System (Applied Biosystems).
<sup>f</sup> Total RNA yield from urine sample 8 was below detection limit for measurement using the Qubit Fluorometer and RNA was insufficient for reverse transcription to cDNA and measurement of 18S rRNA, TGF-β1 mRNA and miR-26a.
<sup>g</sup> P values derived using Wilcoxon matched pairs signed-ranks test.

**Table 2B.** Characteristics of study cohort.

| **Characteristics of Kidney Allograft Recipients** |  |
| --- | --- |
| Number of patients | 12 |
| Age, Years, Median (Min- Max) | 55 (23-77) |
| Sex, Male / Female | 9 / 3 |
| Time from Transplantation to Urine Collection, Months, Median (Min- Max) | 34 (15-137) |
| Type of Kidney Donor, Deceased / Living Related / Living Unrelated | 7 / 3 / 2 |

The absolute 18S rRNA copy number in the 11 urine specimens from 11 unique kidney allograft recipients was 8.18E+08 (3.04E+08 and 1.99 E+09) copies per microgram of RNA with the 11 filtrates stored at -80°C for 2 or 4 days and 1.13E+09 (5.86E+08 and 2.46E+09) copies per microgram of RNA in the paired 11 filtrates stored at ambient temperature for identical duration (P=0.24) (Table 2A).

In addition to measuring absolute copy number of 18S rRNA, we measured absolute copy number of a constitutively expressed mRNA for TGF-β1, a multifunctional cytokine with documented role in transplantation immunity and kidney allograft fibrosis. The absolute TGF-β1 mRNA copy number in the 11 filtrates stored at -80°C was 4.76E+03 (9.77E+02 and 9.11E+03) copies per microgram of RNA and 3.44E+03 (4.64E02+03 and 8.11E+03) copies per microgram of RNA in the paired 11 filtrates stored at ambient temperature for 2 or 4 days (P= 0.08) (Table 2A).

Total RNA isolated using the ZFBP could be predicted to contain miRNAs in addition to rRNAs and mRNAs. We therefore determined miRNA presence in the filtrates by reverse transcribing total RNA using miR-26a specific oligonucleotide primer pair and TaqMan probe and measured absolute copy number using the customized RT-qPCR assay. We selected miR-26a for quantification in view of its role in regulating kidney biology. miR-26a was present in high abundance in the filtrates stored either at -80°C or at ambient temperature for 2 or 4 days. The absolute miR-26a copy number in the 11 filtrates stored at -80°C was 9.17E+05 (6.67E+05 and 1.43E+06) copies per microgram of RNA and 8.34E+05 (6.08E+05 and 1.69E+06) copies per microgram of RNA in the paired 11 filtrates stored at ambient temperature for 2 or 4 days (P=0.58) (Table 2A).

Our findings demonstrate that the three major types of RNA - ribosomal RNA, messenger RNA and microRNA - are all measurable in the total RNA isolated using ZFBP. Furthermore, the findings that RNA purity, abundance and absolute copy numbers of 18S rRNA, TGF-β1 mRNA and miR-26a are similar between the RNA isolated from the filtrate that was stored at - 80°C for 2 or 4 days and the RNA isolated from the paired filtrates stored at ambient temperature for identical duration authenticate RNA stability in the filtrates stored at ambient temperature and lack of a need for a -80°C freezer to process urine specimens.

### 3.3. Urinary Cell Profiling with Filtrates Shipped at Ambient Temperature

We examined the performance characteristics of RNA mailed at ambient temperature to RNA stored at -80°C using 12 urine samples collected from 12 unique kidney allograft recipients. Prior to processing, 5 urine samples from 5 of the 12 recipients were pooled as Batch 1, 4 urine samples from 4 recipients were pooled as Batch 2 and 3 urine samples from 3 recipients were pooled as Batch 3. The urine samples were processed using ZFBP and the filtrates were either stored at -80°C or mailed back to our GEM Core at ambient temperature using a commercial carrier (Figure 3). On receipt of the mailed filtrate the following day, we retrieved the filtrate stored at -80°C in the Core lab and isolated total RNA from the stored filtrate and the mailed filtrate using the Zymo-spin column and analyzed for RNA purity, RNA yield and absolute copy numbers of 18S rRNA, TGF-β1 mRNA and miR-26a.

**Figure 3.**
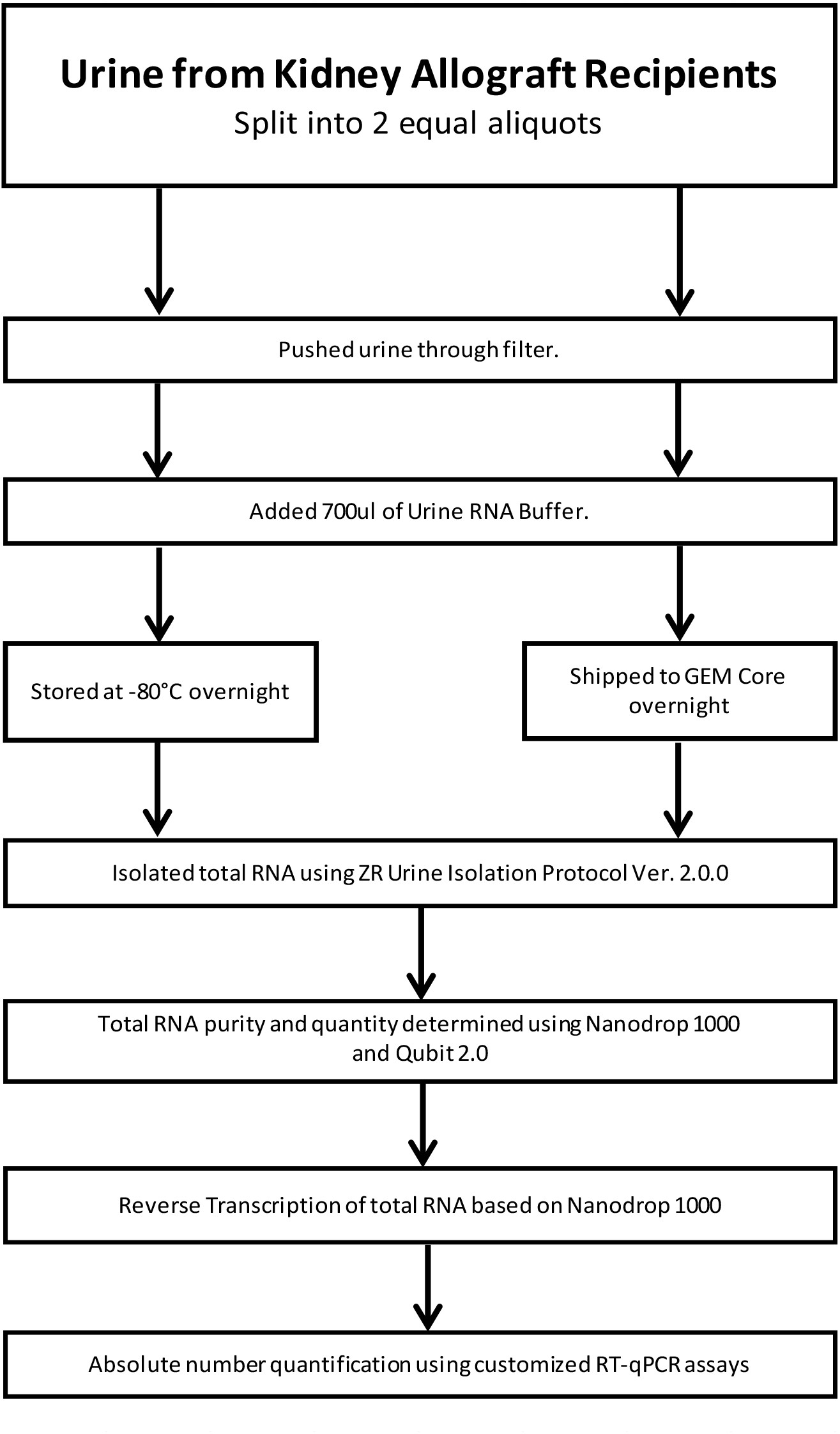
Flow chart for the evaluation of urine filtrates stored either at -80°C or mailed overnight to the GEM Core at ambient temperature. Urine collected from kidney allograft recipients were pooled (Table 3B), divided into 2 equal aliquots, and processed using the ZFBP. In the ZFBP, urine was pushed through the filter and 700μl of urine RNA buffer was added to the ZRC GF™ filter to collect urinary cell filtrate containing total RNA. The filtrates were either stored at -80°C or mailed to GEM Core at ambient temperature. Total RNA was isolated using the ZR Urine RNA Isolation Kit (Zymo, Cat. R1038/1039 Ver.2.0.0) was measured using the Nanodrop 1000 Spectrophotometer and Qubit 2.0 fluorometer. The Nanodrop measurement replicates were within 1ng/μl concentration and 0.2 for the A260/280 ratio. The Qubit readings were measured once using the Qubit RNA BR (Broad-Range) Assay Kit. RNA was reverse transcribed, based on the Nanodrop 1000 readings to cDNA, using TaqMan reverse transcription kit (Applied Biosystems, Cat. N8080234) for mRNA measurement. RNA was reverse transcribed based on the Nanodrop readings using primer-specific TaqMan MicroRNA reverse transcription kit (Applied Biosystems, Cat. 4366597) for miRNA measurement. Absolute copy number of 18S rRNA, TGF-β1 mRNA, and hsa-miR-26a miRNA were measured using gene specific oligonucleotide pairs and TaqMan probes in customized RT-qPCR assays on 7500 Fast Real-Time PCR System

Data from the filtrates either stored at -80°C or mailed back to our GEM Core at ambient temperature using a commercial carrier are shown in Table 3A and the demographics of the study cohort are summarized in Table 3B. The median (25^th^ percentile and 75^th^ percentile) A260/A280 ratio was 2.04 (1.85 and 2.06) for the RNA isolated from the 3 filtrates stored at - 80°C and 2.01 (2.00 and 2.13) for the RNA isolated from the paired filtrates shipped overnight at ambient temperature (P=0.50). The amount of RNA isolated from the 3 filtrates stored at - 80°C was 1.83 (0.83 and 2.94) micrograms and 1.59 (0.79 and 2.57) micrograms from the paired filtrates shipped overnight at ambient temperature (P=0.25). These measurements were made using the NanoDrop spectrophotometer. We also measured RNA yield from the filtrates using the Qubit fluorometer and found that the median amount of total RNA isolated from the 3 filtrates stored at -80°C was 1.01 (0.42 and 1.39) micrograms and 1.07 (0.44 and 1.32) micrograms from the paired filtrates shipped overnight at ambient temperature (P>0.99).

**Table 3A.**
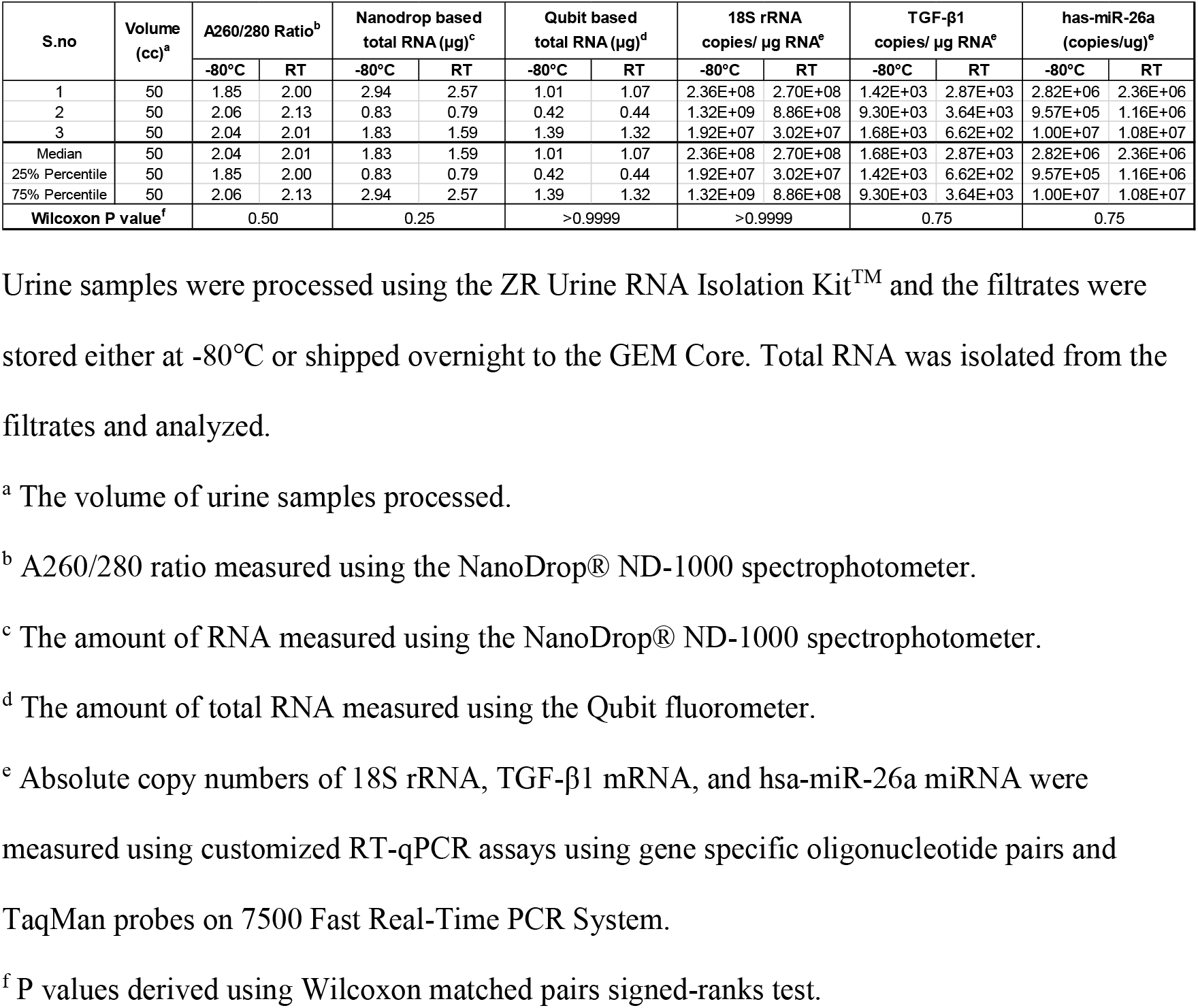
Performance characteristics of RNA isolated from the filtrates stored at -80°C or shipped to the GEM Core at Ambient (Room) Temperature.

**Table 3B.** Characteristics of study cohort.

| Characteristics of Kidney Allograft Recipients |  |
| --- | --- |
| Number of Patients |  |
| Batch 1 | 5 |
| Batch 2 | 4 |
| Batch 3 | 3 |
| Age, Years, Median (Min- Max) |  |
| Batch 1 | 57 (25-74) |
| Batch 2 | 54 (33-60) |
| Batch 3 | 53 (38-81) |
| Gender, Male / Female |  |
| Batch 1 | 4 / 1 |
| Batch 2 | 1 / 3 |
| Batch 3 | 2 / 1 |
| Time from Transplantation to Urine Collection, Months, Median (Min- Max) |  |
| Batch 1 | 36 (11-127) |
| Batch 2 | 34 (17-84) |
| Batch 3 | 33 (29-73) |
| Type of Transplant Donor, Deceased / Living Related / Living Unrelated |  |
| Batch 1 | 2 / 2 / 1 |
| Batch 2 | 3 / 0 / 1 |
| Batch 3 | 3 / 0 / 0 |

The absolute copy number of 18S rRNA was 2.36E+08 (1.92E+07 and 1.32E+09) per microgram of total RNA in the filtrates stored at -80°C and 2.70 E+08 (3.02E+07 and 8.86E+08) in the paired filtrate shipped overnight at ambient temperature (P>0.99). The absolute copy number of TGF-β1 mRNA was 1.68E+03 (1.42E+03 and 9.30E+03) in the filtrates stored at -80°C and 2.87E+03 (6.62E+02 and 3.64E+03) in the paired filtrate shipped overnight at ambient temperature (P=0.75). The absolute copy number of miR-26a copy number was 2.82E+06 (9.57E+05 and 1.00E+07) in the filtrates stored at -80°C and 2.36E+06 (1.16E+06 and 1.08E+07) in the paired filtrates shipped overnight at ambient temperature (P=0.75).

Our findings that RNA purity, amount, and absolute copy numbers of 18s rRNA, TGF-β1 mRNA and miR-26a are similar between the filtrates stored at -80°C or shipped overnight at ambient temperature demonstrate the feasibility of shipment of filtrates containing RNA at ambient temperature.

### 3.4. Urinary Cell Profiling of Filtrates Processed at Home by Kidney Allograft Recipients

Home processing of urine samples by kidney transplant recipients has several advantages including obviating the need for visits to a laboratory or a clinic for urine collection. We therefore examined the feasibility of initial processing of urine by the patients (Figure 4). The kidney transplant recipients were trained by GEM Core staff, during their scheduled visits to the transplant clinic, to push urine through the filter and then collect the filtrate containing RNA by pushing the RNA buffer through the filter. Following their training, the patients were asked to collect the filtrates at home one day prior to their scheduled clinic visit and bring the filtrates to the clinic. A fresh urine sample was also collected at the time of their clinic visit and this urine sample was processed by the GEM Core staff.

**Figure 4.**
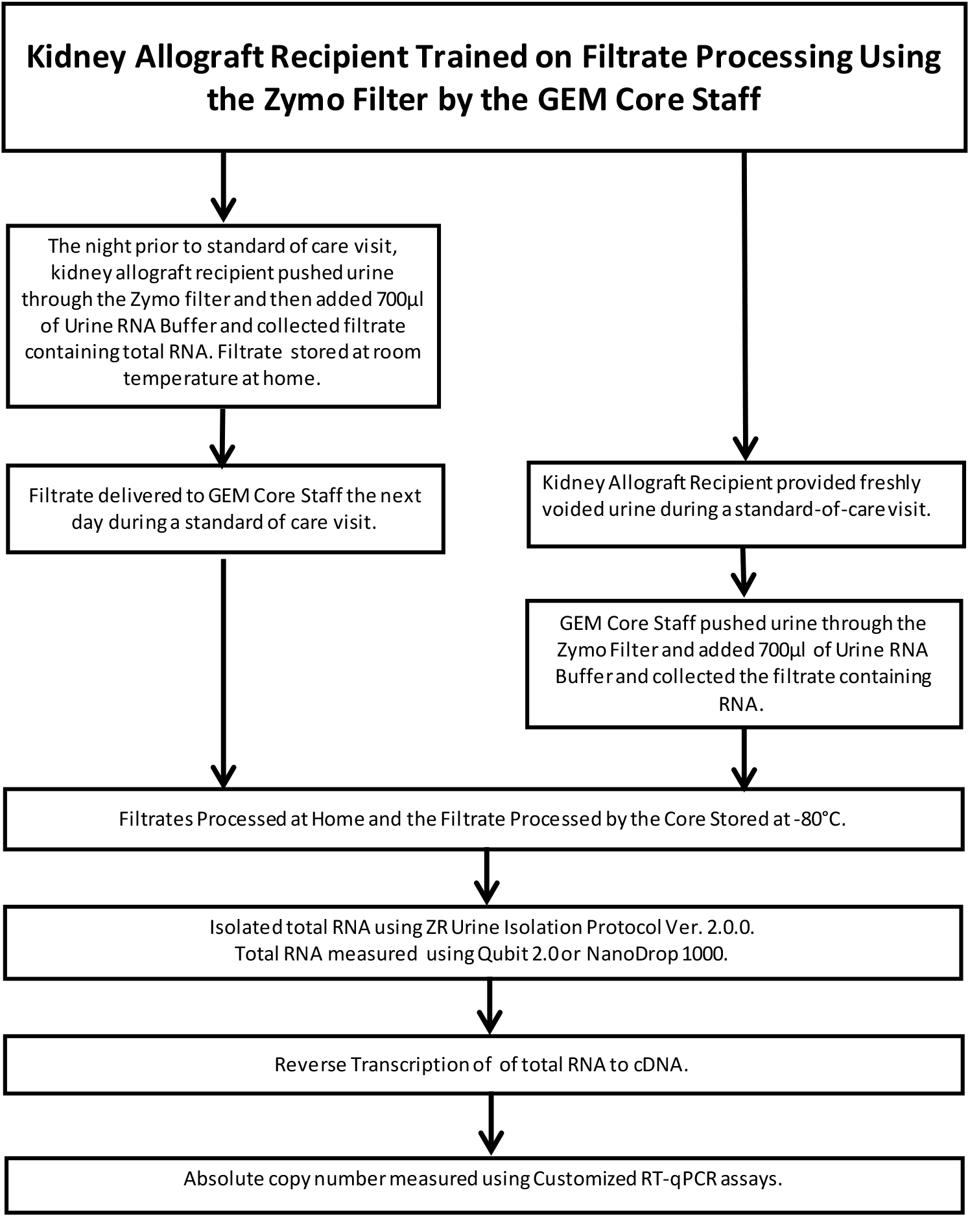
Flow chart for assessing filtrates processed at home by the kidney allograft recipients and filtrates processed in the GEM Core. Kidney allograft recipients were trained by the Gene Expression Monitoring (GEM) Core staff to push urine through the Zymo filter followed by pushing of 700μL of Urine RNA Buffer and collect the filtrate containing total RNA. A urine processing kit containing all necessary materials was provided to each patient to perform the initial processing steps at home. Patients were instructed to collect the filtrate the night before they were to come in for their standard-of-care visit to the transplant clinic. The home collected filtrate was left at ambient temperature and brought to the clinic the following day. The home collected filtrate and a freshly voided urine sample were provided to the GEM Core staff. The GEM Core staff processed the fresh urine using the ZR Urine RNA Isolation Kit. The home processed filtrate and the Core processed filtrate were stored at -80°C prior to RNA isolation. Total RNA was isolated using the Zymo-Spin™ IC columns and the isolated RNA was measured using the Nanodrop 1000 Spectrophotometer and the Qubit 2.0 fluorometer. The Nanodrop measurement replicates were within 1ng/μl concentration and 0.2 for the A260/280 ratio. The Qubit readings were measured once using the Qubit RNA BR (Broad-Range) Assay Kit. RNA was reverse transcribed to cDNA based on Qubit Assay readings. In one patient, Nanodrop® 1000 readings were used for the reverse transcription reaction. RNA was reverse transcribed to cDNA using TaqMan reverse transcription kit. Absolute copy numbers of 18S rRNA and TGF-β1 mRNA were measured using gene specific primer pairs and gene specific TaqMan probes in the customized RT-qPCR assays on a 7500 Fast Real-Time PCR System.

We compared the performance characteristics of RNA isolated from the home processed filtrates with the RNA isolated by the GEM Core staff. A total of 25 urine specimens were processed at home by 5 kidney allograft recipients (Table 4A and 4B). Among these samples, 3 urine specimens from a single patient did not yield sufficient RNA for reverse transcription and these samples were excluded from downstream analysis. Among the remaining 22 urine samples, the median (25^th^ percentile and 75^th^ percentile) A260/A280 ratio for the RNA isolated from the 22 filtrates collected at home by the patients was 1.97 (1.88 and 2.02) and 1.99 (1.93 and 2.02) for the RNA isolated by the GEM Core Staff (Mann Whitney P=0.57). Total RNA yield from the home processed urine samples was 1.66 (0.76 and 3.16) micrograms and 2.12 (0.84. and 5.05) micrograms for the Core processed samples (P=0.75), as measured using the NanoDrop spectrophotometer. Total RNA yield, as measured using Qubit fluorometer, was 0.89 (0.46 and 2.74) micrograms and 1.57 (0.60 and 3.94) micrograms for the Core processed samples (P=0.47). The RNA yield from the home processed samples and the Core processed samples are not comparable since the volume of urine processed at home and at the Core were not similar and in 5 instances the volume of urine processed at home were not recorded.

**Table 4A.**
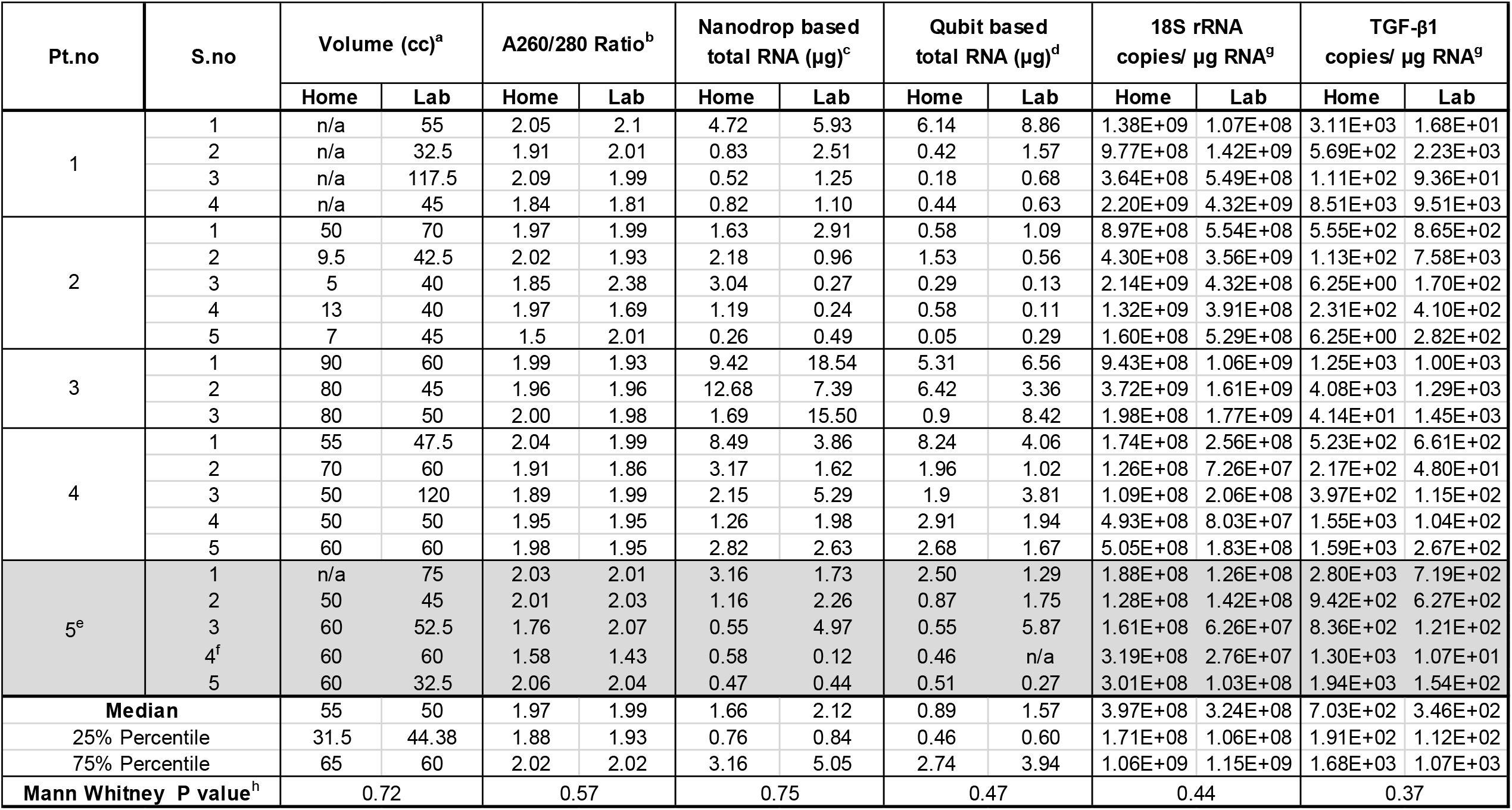
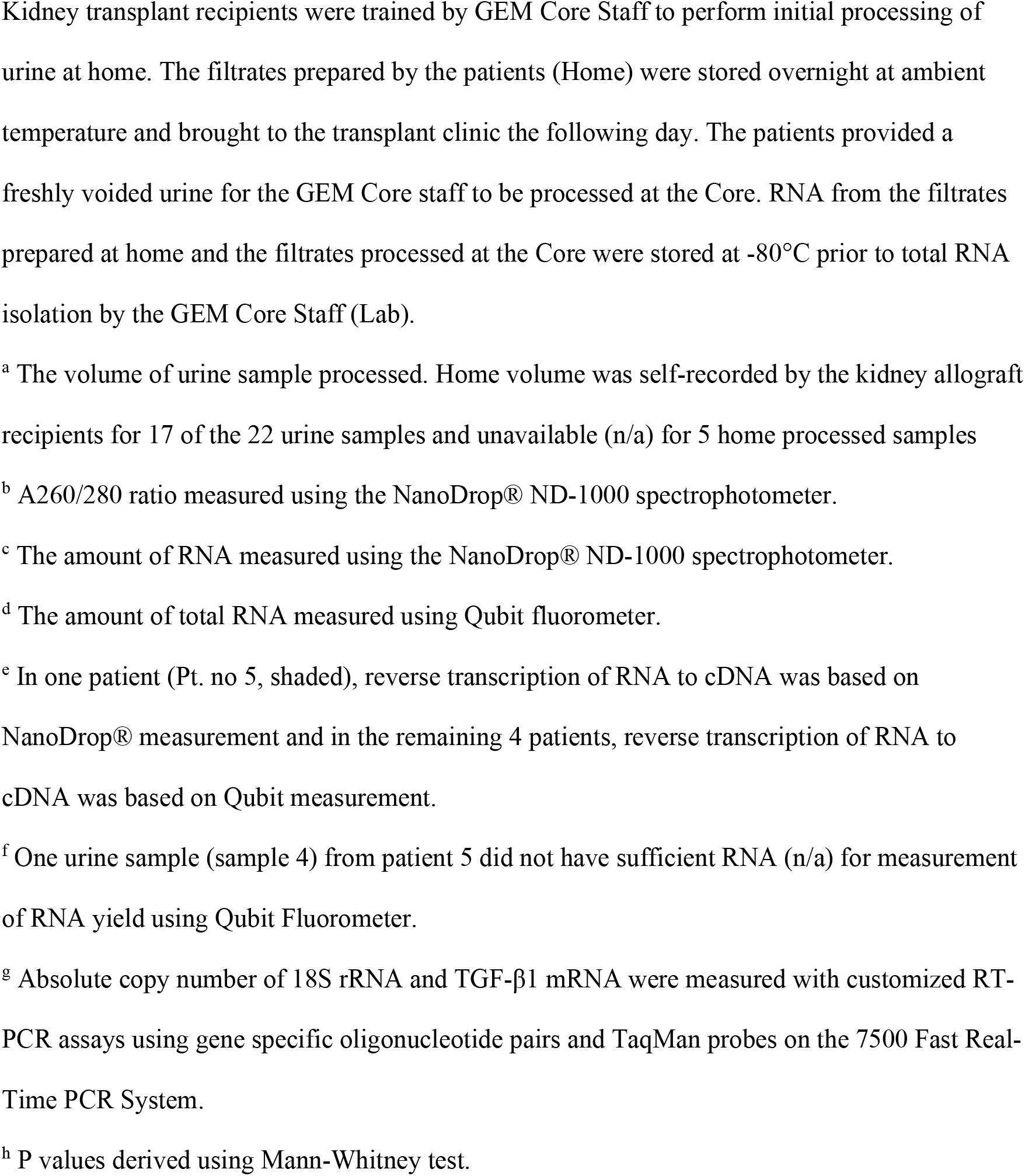
Performance characteristics of RNA isolated from home processed filtrates vs. GEM Core processed filtrates.

| Pt.no | S.no | Volume (cc)<sup>a</sup> |  | A260/280 Ratio<sup>b</sup> |  | Nanodrop based total RNA (µg)<sup>c</sup> |  | Qubit based total RNA (µg)<sup>d</sup> |  | 18S rRNA copies/ µg RNA<sup>g</sup> |  | TGF-β1 copies/ µg RNA<sup>g</sup> |  |
| --- | --- | --- | --- | --- | --- | --- | --- | --- | --- | --- | --- | --- | --- |
|  |  | Home | Lab | Home | Lab | Home | Lab | Home | Lab | Home | Lab | Home | Lab |
| 1 | 1 | n/a | 55 | 2.05 | 2.1 | 4.72 | 5.93 | 6.14 | 8.86 | 1.38E+09 | 1.07E+08 | 3.11E+03 | 1.68E+01 |
|  | 2 | n/a | 32.5 | 1.91 | 2.01 | 0.83 | 2.51 | 0.42 | 1.57 | 9.77E+08 | 1.42E+09 | 5.69E+02 | 2.23E+03 |
|  | 3 | n/a | 117.5 | 2.09 | 1.99 | 0.52 | 1.25 | 0.18 | 0.68 | 3.64E+08 | 5.49E+08 | 1.11E+02 | 9.36E+01 |
|  | 4 | n/a | 45 | 1.84 | 1.81 | 0.82 | 1.10 | 0.44 | 0.63 | 2.20E+09 | 4.32E+09 | 8.51E+03 | 9.51E+03 |
| 2 | 1 | 50 | 70 | 1.97 | 1.99 | 1.63 | 2.91 | 0.58 | 1.09 | 8.97E+08 | 5.54E+08 | 5.55E+02 | 8.65E+02 |
|  | 2 | 9.5 | 42.5 | 2.02 | 1.93 | 2.18 | 0.96 | 1.53 | 0.56 | 4.30E+08 | 3.56E+09 | 1.13E+02 | 7.58E+03 |
|  | 3 | 5 | 40 | 1.85 | 2.38 | 3.04 | 0.27 | 0.29 | 0.13 | 2.14E+09 | 4.32E+08 | 6.25E+00 | 1.70E+02 |
|  | 4 | 13 | 40 | 1.97 | 1.69 | 1.19 | 0.24 | 0.58 | 0.11 | 1.32E+09 | 3.91E+08 | 2.31E+02 | 4.10E+02 |
|  | 5 | 7 | 45 | 1.5 | 2.01 | 0.26 | 0.49 | 0.05 | 0.29 | 1.60E+08 | 5.29E+08 | 6.25E+00 | 2.82E+02 |
| 3 | 1 | 90 | 60 | 1.99 | 1.93 | 9.42 | 18.54 | 5.31 | 6.56 | 9.43E+08 | 1.06E+09 | 1.25E+03 | 1.00E+03 |
|  | 2 | 80 | 45 | 1.96 | 1.96 | 12.68 | 7.39 | 6.42 | 3.36 | 3.72E+09 | 1.61E+09 | 4.08E+03 | 1.29E+03 |
|  | 3 | 80 | 50 | 2.00 | 1.98 | 1.69 | 15.50 | 0.9 | 8.42 | 1.98E+08 | 1.77E+09 | 4.14E+01 | 1.45E+03 |
| 4 | 1 | 55 | 47.5 | 2.04 | 1.99 | 8.49 | 3.86 | 8.24 | 4.06 | 1.74E+08 | 2.56E+08 | 5.23E+02 | 6.61E+02 |
|  | 2 | 70 | 60 | 1.91 | 1.86 | 3.17 | 1.62 | 1.96 | 1.02 | 1.26E+08 | 7.26E+07 | 2.17E+02 | 4.80E+01 |
|  | 3 | 50 | 120 | 1.89 | 1.99 | 2.15 | 5.29 | 1.9 | 3.81 | 1.09E+08 | 2.06E+08 | 3.97E+02 | 1.15E+02 |
|  | 4 | 50 | 50 | 1.95 | 1.95 | 1.26 | 1.98 | 2.91 | 1.94 | 4.93E+08 | 8.03E+07 | 1.55E+03 | 1.04E+02 |
|  | 5 | 60 | 60 | 1.98 | 1.95 | 2.82 | 2.63 | 2.68 | 1.67 | 5.05E+08 | 1.83E+08 | 1.59E+03 | 2.67E+02 |
| 5<sup>e</sup> | 1 | n/a | 75 | 2.03 | 2.01 | 3.16 | 1.73 | 2.50 | 1.29 | 1.88E+08 | 1.26E+08 | 2.80E+03 | 7.19E+02 |
|  | 2 | 50 | 45 | 2.01 | 2.03 | 1.16 | 2.26 | 0.87 | 1.75 | 1.28E+08 | 1.42E+08 | 9.42E+02 | 6.27E+02 |
|  | 3 | 60 | 52.5 | 1.76 | 2.07 | 0.55 | 4.97 | 0.55 | 5.87 | 1.61E+08 | 6.26E+07 | 8.36E+02 | 1.21E+02 |
|  | 4<sup>f</sup> | 60 | 60 | 1.58 | 1.43 | 0.58 | 0.12 | 0.46 | n/a | 3.19E+08 | 2.76E+07 | 1.30E+03 | 1.07E+01 |
|  | 5 | 60 | 32.5 | 2.06 | 2.04 | 0.47 | 0.44 | 0.51 | 0.27 | 3.01E+08 | 1.03E+08 | 1.94E+03 | 1.54E+02 |
| Median |  | 55 | 50 | 1.97 | 1.99 | 1.66 | 2.12 | 0.89 | 1.57 | 3.97E+08 | 3.24E+08 | 7.03E+02 | 3.46E+02 |
| 25% Percentile |  | 31.5 | 44.38 | 1.88 | 1.93 | 0.76 | 0.84 | 0.46 | 0.60 | 1.71E+08 | 1.06E+08 | 1.91E+02 | 1.12E+02 |
| 75% Percentile |  | 65 | 60 | 2.02 | 2.02 | 3.16 | 5.05 | 2.74 | 3.94 | 1.06E+09 | 1.15E+09 | 1.68E+03 | 1.07E+03 |
| Mann Whitney P value<sup>h</sup> |  | 0.72 |  | 0.57 |  | 0.75 |  | 0.47 |  | 0.44 |  | 0.37 |  |

Kidney transplant recipients were trained by GEM Core Staff to perform initial processing of urine at home. The filtrates prepared by the patients (Home) were stored overnight at ambient temperature and brought to the transplant clinic the following day. The patients provided a freshly voided urine for the GEM Core staff to be processed at the Core. RNA from the filtrates prepared at home and the filtrates processed at the Core were stored at -80°C prior to total RNA isolation by the GEM Core Staff (Lab).
<sup>a</sup> The volume of urine sample processed. Home volume was self-recorded by the kidney allograft recipients for 17 of the 22 urine samples and unavailable (n/a) for 5 home processed samples
<sup>b</sup> A260/280 ratio measured using the NanoDrop® ND-1000 spectrophotometer.
<sup>c</sup> The amount of RNA measured using the NanoDrop® ND-1000 spectrophotometer.
<sup>d</sup> The amount of total RNA measured using Qubit fluorometer.
<sup>e</sup> In one patient (Pt. no 5, shaded), reverse transcription of RNA to cDNA was based on NanoDrop® measurement and in the remaining 4 patients, reverse transcription of RNA to cDNA was based on Qubit measurement.
<sup>f</sup> One urine sample (sample 4) from patient 5 did not have sufficient RNA (n/a) for measurement of RNA yield using Qubit Fluorometer.
<sup>g</sup> Absolute copy number of 18S rRNA and TGF-β1 mRNA were measured with customized RT-PCR assays using gene specific oligonucleotide pairs and TaqMan probes on the 7500 Fast Real-Time PCR System.
<sup>h</sup> P values derived using Mann-Whitney test.

**Table 4B.** Characteristics of study cohort.

| **Characteristics of Kidney Allograft Recipients** |  |
| --- | --- |
| Number of Patients | 5 |
| Age, Years, Median (Min- Max) | 47 (33-62) |
| Sex, Male / Female | 2 / 3 |
| Time from Transplantation to Urine Collection, Days, Median (Min- Max) | 32 (11-97) |
| Type of Kidney Donor, Deceased / Living Related / Living Unrelated | 4 / 1 / 0 |

Absolute copy number of 18S rRNA was higher than the 18S rRNA adequacy threshold of 5×10^7^ copies per microgram in all 22 home processed samples and in 21 of 22 Core processed samples, and absolute copy number of 18S rRNA in the 22 home processed samples was 3.97E+08 (1.71E+08 and 1.06E+09) per microgram of RNA and 3.24E+08 (1.06E+08 and 1.15+09) per microgram of RNA in the Core processed samples (P=0.44). Absolute copy number of TGF-β1 mRNA was higher than the TGF-β1 mRNA adequacy threshold of 1×10^2^ copies per microgram in 19 of 22 home processed samples and in 18 of 22 Core processed samples. The absolute copy number of TGF-β1 mRNA in the home processed samples was 7.03E+02 (1.91E+02 and 1.68E+03) per microgram of RNA and 3.46E+02 (1.12E+02 and 1.07E+03) per microgram of RNA in Core processed samples (P=0.37).

Our findings that RNA yield, RNA purity and absolute copy numbers of 18S rRNA and TGF-β1 are similar between urine samples processed at home by kidney transplant recipients and urine samples processed by GEM Core staff demonstrate the feasibility of initial processing of urine samples by the kidney transplant recipients.

### 3.5. Noninvasive Diagnosis of Acute Rejection in Kidney Allografts

Our single center studies and CTOT-4 multicenter study utilized the CCBP to isolate total RNA for urinary cell mRNA profiling, and for the discovery of mRNAs associated with acute rejection (AR) in human kidney allografts (6-18). We examined whether urinary cell mRNA profiling of RNA isolated using the ZFBP is diagnostic of AR.

We collected a total of 22 urine samples matched to kidney allograft biopsies from 20 unique kidney allograft recipients. Among the 22 samples, 8 were matched to AR biopsies and 14 were matched to biopsies without acute or chronic rejection (NR biopsies). Among the 8 AR biopsy matched urine samples, urine volume was 7.5 ml for one sample and TGF-β1 mRNA copy number was less than 100 copies per microgram of RNA in another sample, and these two samples were excluded from downstream analysis. Among the 14 NR biopsy matched urine samples, urine volume was 10 ml in one sample and TGF-β1 mRNA copy number was less than 100 copies per microgram of RNA in two samples and these three samples were also excluded from downstream analysis.

The AR biopsies and NR biopsies were categorized using the Banff biopsy classification schema (21). Among the 6 AR biopsies from 6 unique kidney allograft recipients (Age 31-62 years; 5 males and 1 female; 2 deceased donor kidney allografts and 4 living donor kidney allografts), 3 biopsies were classified as ACR biopsies and the remaining 3 were classified as both ACR and AMR. Among the 11 NR biopsies from 10 unique kidney allograft recipients (Age 31-74 years; 5 males and 5 females; 7 deceased donor kidney allografts and 3 living donor kidney allografts), 3 biopsies showed features of acute tubular necrosis (ATN), 3 were classified as diabetic nephropathy, 2 as focal and segmental sclerosis (FSGS), one as interstitial fibrosis and tubular atrophy (IFTA) biopsy and 2 as normal biopsies.

The absolute copy number of the reference gene 18S rRNA was 2.3E+09 (1.5E+09 and 2.9 E+09) copies per microgram of RNA in the urine matched to AR biopsies and 1.93E+09 (7.16E+08 and 3.76E+09) copies per microgram in the urine matched to NR biopsies and urinary cell levels of 18S rRNA did not discriminate AR biopsies from NR biopsies (P=1.00) (Figure 5A). The absolute copy number of the constitutively expressed TGF-β1 mRNA was 2760 (1300 and 35675) copies per microgram of RNA in the urine matched to AR biopsies and 4150 (703 and 22500) copies per microgram of RNA in the urine matched to NR biopsies and urinary cell levels of TGF-β1 mRNA did not discriminate AR biopsies from NR biopsies (P=0.81) (Figure 5B). In accord with previous studies (8, 15), urinary cell levels of mRNA for granzyme B and perforin were significantly higher in urine matched to AR biopsies compared to urine matched to NR biopsies. The absolute copy number of granzyme B mRNA was 7415 (1790 and 21400) copies per microgram of RNA in the urine matched to AR biopsies and was significantly higher than the 694 (187 and 2920) copies per microgram of RNA in the urine matched to NR biopsies (P=0.01) (Fig. 5C). The absolute copy number of perforin mRNA was 1029 (534 and 2981) copies per microgram of RNA in the urine matched to AR biopsies and 111 (50 and 235) copies per microgram of RNA in the urine matched to NR biopsies (P=0.00002) (Fig. 5D). Altogether, our data demonstrate that noninvasive diagnosis of acute rejection is feasible using ZFBP to process urine from kidney allograft recipients.

**Figure 5.**
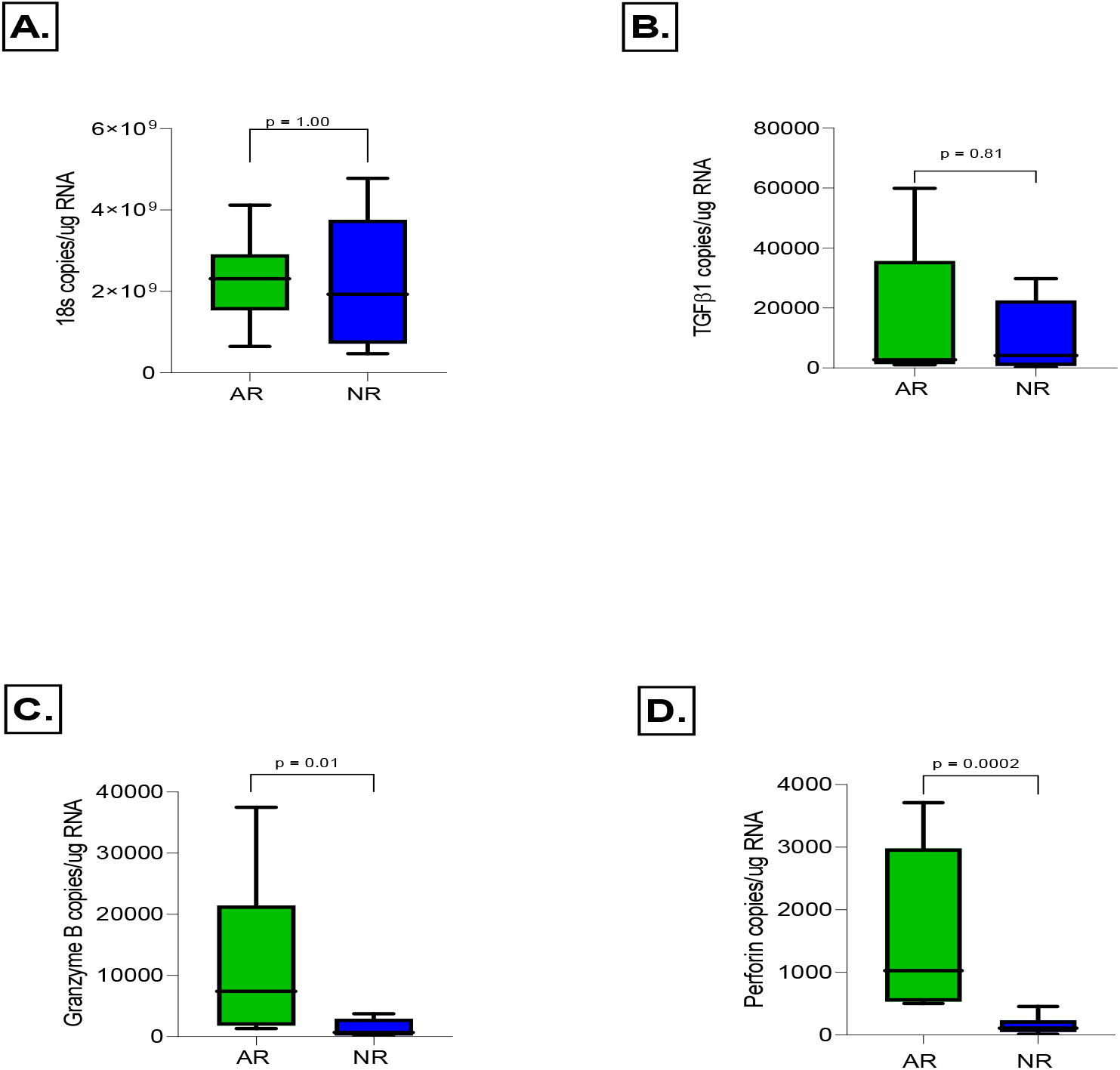
Levels of urinary cell transcripts. Box and whisker plots show the 10^th^, 25^th^, 50^th^ (median), 75^th^ and 90^th^ percentiles values for 18S rRNA (A), TGF-β1 mRNA (B), granzyme B mRNA (C) and perforin mRNA (D) in biopsy matched urine samples from kidney allograft recipients with kidney allograft biopsies showing acute rejection (AR, n=6) or biopsies without histological features of acute or chronic rejection (NR, n=11). The levels of granzyme B mRNA (P=0.01) and perforin mRNA (P=0.0002), but not the levels of 18S rRNA (P=1.00) or TGF-β1 mRNA (P=0.81) were significantly higher in urine matched to AR biopsies compared to urine matched to NR biopsies. P values calculated using Mann-Whitney test.

## 4. DISCUSSION

We investigated a filter-based protocol for the processing of urine samples for urinary cell mRNA profiling of kidney allograft recipients and demonstrate the following distinct advantages over the conventional centrifugation-based protocol: (i) the kidney transplant patients themselves can perform the initial processing steps and there is no need for a centrifuge to prepare urine cell pellets; (ii) RNA in the filtrates is stable at ambient temperature and -80°C storage of urinary cell pellets prior to shipping to a central laboratory can be altogether eliminated; and (iii) shipping at ambient temperature to the laboratory for further processing is also feasible and special packaging of samples for shipping is not necessary. Because the patients themselves can perform the initial steps up to the collection of filtrates containing RNA, the need for prompt processing of collected urine samples by laboratory staff is eliminated. The elimination of specialized equipment such as centrifuges and -80°C freezers, and the stability of the RNA containing filtrate at ambient temperature represent significant advantages. The ZFBP examined in this study however requires the patients to perform multiple tasks, albeit simple ones such as pushing the urine and the RNA buffer sequentially through the filter and collecting the filtrate. Development of an automated mechanical device could make initial processing even easier.

A number of earlier studies have evaluated filtration-based methods to isolate RNA from urinary cells (19-20), but none, to our knowledge, evaluated patient’s ability to perform the initial steps of urine processing. We also demonstrate measurement of absolute copy number of all three types of RNAs-18S ribosomal RNA, mRNA and microRNA. Importantly, noninvasive diagnosis of AR was accomplished with urine samples processed using the ZFBP.

A large body of data demonstrate that urinary cell levels of mRNA encoding immunoregulatory proteins are diagnostic of acute rejection in human kidney allografts (6-16). All of these studies utilized the centrifugation-based protocols to sediment urinary cells. Our demonstration that RNA purity and 18S rRNA copy numbers are similar if not better with the ZFBP compared to CCBP (Table 1) suggests that the filtration-based protocol is a suitable alternative to the centrifugation-based protocol.

We examined a number of issues of significance to the clinical use of ZFBP. We identified report that the RNA in the filtrate collected as flow-through is stable at ambient temperature for 2 or 4 days (Table 2) and can be shipped at ambient temperature (Table 3). These findings eliminate the need for prompt processing of urine samples and the refrigeration of urine cell pellets for storage or shipment.

A novel and clinically relevant observation from this study is the demonstration that kidney allograft recipients could be trained to perform the initial processing of urine at home (Table 4). Clearly, our proof-of-principle study needs to be expanded to include a larger cohort, but we anticipate no significant challenges in widespread implementation in view of kidney transplant patient’s ability to maintain records and follow complex post-transplant medication regimens. Development of an automated mechanical device, similar to a continuous glucose monitoring device, would make the process even easier for the patients.

To our knowledge, this is the first report of noninvasive diagnosis of acute rejection using a filtration-based protocol to isolate RNA from urinary cells. The finding that there is almost a 10-fold difference in the copy numbers of granzyme B and perforin in urine matched to acute rejection biopsies compared to urine matched to biopsies without rejection features (Figure 5) parallels the differences in the abundances of perforin and granzyme B we previously identified using the centrifugation method (15). It will be important however to perform a study involving a larger cohort of kidney allograft recipients.

In sum, we have investigated a ZFBP for urinary cell mRNA profiling of kidney allograft recipients and identified that RNA yield, purity and the abundance of transcripts in the isolated RNA are similar, if not better, than the performance characteristics of RNA isolated using the conventional centrifugation-based method. The feasibility and the accuracy of ZFBP to process urine specimens should enable wider implementation of urinary cell mRNA profiling.

## Data Availability

Data available in manuscript. No additional external datasets or supplementary material.

## Acknowledgements

The authors gratefully acknowledge the exceptional contributions of our colleagues Drs. Phyllis August, Darshana Dadhania, John Lee and Thangamani Muthukumar.

